# Expression of specific *var* gene subtypes is associated with distinct severe malaria syndromes

**DOI:** 10.64898/2026.02.02.26345432

**Authors:** Henry Ndugwa, Michelle Muthui, J. Alexandra Rowe, SM Kinyanjui, Cheryl Andisi Kivisi, Abdirahman I. Abdi

## Abstract

The virulence of *Plasmodium falciparum* is closely linked to *P. falciparum* erythrocyte membrane protein 1 (*Pf*EMP1), encoded by the diverse *var* gene family. *Pf*EMP1 mediates parasite immune evasion and vascular adhesion of infected red blood cells, contributing to severe disease. While expression of group A or domain cassette (DC8 and DC13)-containing *var* genes have been associated with severe malaria, direct comparisons between the severe malaria syndromes remain limited. Furthermore, interactions between specific *var* gene expression and the rosetting phenotype, a known marker of severe malaria, are incompletely understood across severe malaria syndromes.

We analysed parasite and clinical data from 966 Kenyan children with non-severe malaria and severe malaria syndromes collected over 18 years in Kilifi, Kenya. *Var* gene transcription was profiled using DBLα-tag sequencing (n=898), RT-qPCR (n=768), and DBLα-tag-based prediction tools.

We show that parasites expressing group A or DC8 *var* genes were associated with impaired consciousness (IC), while rosetting was associated with respiratory distress (RD) and severe malarial anaemia (SMA), but not IC. Variant expression was more homogeneous in SMA than IC or RD. These findings suggest that IC, RD and SMA reflect distinct underlying parasite virulence strategies: sequestration, rosetting-associated parasite virulence, and chronicity of infection respectively.

In conclusion, this analysis provides the most comprehensive characterisation of *P. falciparum var* gene expression across the major severe malaria syndromes to date and highlights the possibility of syndrome-specific parasite virulence mechanisms that may inform the development of targeted therapeutic interventions

**Graphical abstract:** 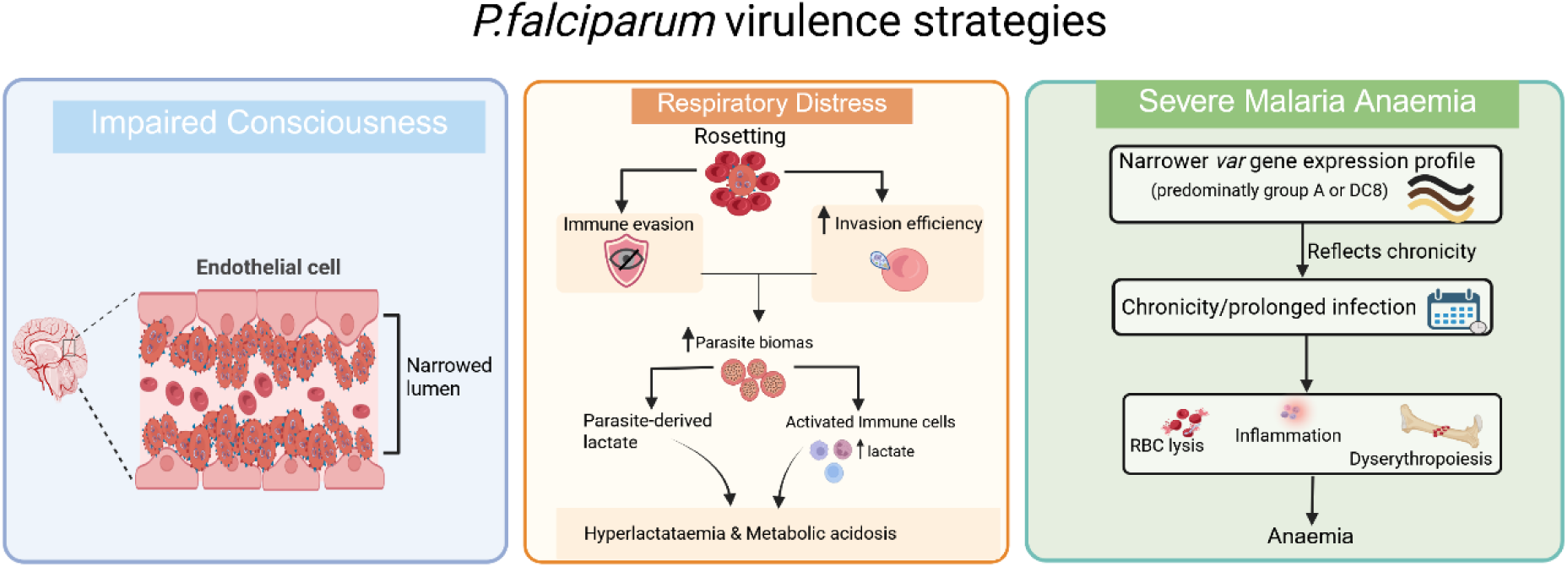

## Introduction

*Plasmodium falciparum* malaria remains a major global health burden, causing over half a million deaths annually, primarily in sub-Saharan Africa (WHO, 2023). The outcome spectrum of malaria infection ranges from asymptomatic through non-severe to severe malaria, with severe malaria presenting with life-threatening complications such as impaired consciousness (IC), severe malarial anaemia (SMA), and respiratory distress (RD) (Marsh et al., 1995). A deeper understanding of host-parasite interactions at the molecular level that drive distinct clinical malaria syndromes is essential for the development of targeted therapies. A key unresolved question in malaria research is whether parasites that cause severe disease differ biologically from those responsible for non-severe malaria (NSM), and whether specific severe manifestations can be attributed to particular parasite features.

Central to the pathogenesis of severe malaria is the adhesion of infected red blood cells (iRBCs) to endothelial cells in the microvasculature, a process known as cytoadhesion. Cytoadhesion facilitates immune evasion by preventing spleen-mediated clearance of iRBCs (Saul, 1999; Rowe et al., 2009). The primary parasite ligands mediating these interactions are the *Plasmodium falciparum* erythrocyte membrane protein 1 (*Pf*EMP1) family. *Pf*EMP1 proteins mediate cytoadhesion by binding to endothelial receptors such as endothelial protein C receptor (EPCR), intercellular adhesion molecule-1 (ICAM-1), cluster of differentiation 36 (CD36), complement receptor 1 (CR1), and chondroitin sulfate A (CSA) (Baruch et al., 1996; Rowe et al., 2009; Smith et al., 2013; Turner et al., 2013; Bernabeu et al., 2016; Ndam et al., 2017), with different binding specificities linked to distinct severe malaria phenotypes. For instance, the binding of iRBCs expressing VAR2CSA to CSA on placental syncytiotrophoblasts contributes significantly to placental malaria (Achur et al., 2000; Clausen et al., 2012). Additionally, rosetting, in which iRBCs bind uninfected RBCs, plays a role in severe malaria pathogenesis (Carlson et al., 1990; Kaul et al., 1991; Cockburn et al., 2003; Doumbo et al., 2009).

*Pf*EMP1 is a large (200 to 400-kDa) clonally variant surface antigen encoded by the highly polymorphic *var* gene family (Smith et al., 1995; Gardner et al., 2002). Each *P. falciparum* genome carries approximately 60 *var* genes that are transcribed in a mutually exclusive fashion. This allows the parasite to alter antigenic profiles and evade host immunity (Deitsch & Dzikowski, 2017). *Pf*EMP1 typically includes an N-terminal “head structure” with an N-terminal sequence (NTS) and a Duffy Binding-Like (DBL) domain, followed by a Cysteine-rich InterDomain Region (CIDR) (Smith et al., 2000). *Var* genes are classified into three major groups (A, B, and C) based on their chromosomal location and promoter sequence (Smith, 2014). This classification is clinically relevant, as group A *var* genes are typically associated with severe malaria, whereas group B and C are more frequently expressed during mild or asymptomatic infections (Kyriacou et al., 2006; Rottmann et al., 2006). Additionally, *var* genes are grouped according to conserved domain arrangements known as domain cassettes (DC) (Rask et al., 2010), with parasites expressing DC8 and DC13 strongly correlated with severe disease presentations (Lavstsen et al., 2012).

Further categorisation is based on cysteine residue counts and conserved amino acid motifs (“MFK” and “REY”) at positions of limited sequence variation (PoLV) within DBLα-tags referred to as cp grouping, with six distinct subgroups (cp1-cp6) (Bull et al., 2005, 2007). A complementary classification is based on DBLα-tag sequences that share position-specific polymorphic sequence blocks known as block-sharing group (bs1 and bs2).bs1 sequences largely belong to group A and bs1 sequences containing two cysteine residues are known as bs1_cys2 and are referred to as group A-like (Warimwe et al., 2009). Among these, cp1 and group A-like are linked to IC, whereas cp2 is predominantly associated with SMA (Warimwe et al., 2009).

These findings suggest that parasites expressing certain *Pf*EMP1 variants are more virulent. Despite these advances, previous studies have been limited by small sample sizes and uneven representation of clinical syndromes, which constrain statistical power and generalisability. Kilifi, a coastal region in Kenya has experienced major decline in transmission and shift in the clinical spectrum of malaria in the past two decades (Mogeni et al., 2016; Njuguna et al., 2019; Kamau et al., 2026). This setting provides an opportunity to revisit these questions across well-represented syndromes using samples collected over 2 decades.

Furthermore, previous studies have predominantly measured *var* gene expression using either RT-qPCR targeting specific *var* gene subtypes or sequencing of expressed DBLα-tags in parasite isolates (Bull et al., 2005; Lavstsen et al., 2012; Mkumbaye et al., 2017; Andisi & Abdi, 2022). Each approach has strengths and limitations. RT-qPCR offers quantitative resolution for predefined targets but cannot capture the full *var* repertoire. DBLα-tag sequencing on the other hand, captures *var* repertoire breadth and diversity but lacks precise quantitation of individual transcripts. Reliance on a single method has likely contributed to inconsistent associations with clinical phenotypes reported in previous studies.

To overcome these limitations, this study analysed clinical data, and parasite isolates from 966 children recruited in Kilifi between 1994-2012, representing the spectrum of clinical malaria presentations. Parasite *var* gene expression profiles were determined using both DBLα-tag sequencing and RT-qPCR and these complementary datasets were analysed in parallel to refine the associations between specific *Pf*EMP1 variants and distinct severe malaria syndromes. Unlike previous studies, which primarily compared severe and uncomplicated malaria, this study directly contrasts parasites from IC, RD, SMA and non-severe malaria within a single large cohort, integrating *var gene* expression, rosetting and infected erythrocyte antibody recognition data.

## Results

### Participants and sample characteristics

We analysed clinical data and parasite samples from 966 children with microscopy-confirmed *Plasmodium falciparum* malaria recruited in Kilifi, Kenya, between 1994 and 2012. This period spans three phases of malaria transmission in Kilifi County: high transmission (pre-decline, 1994-2002), declining transmission (decline, 2003-2008), and low transmission (post-decline, 2009-2012) (**Figure S1**) (Mogeni et al., 2016). Of the 966 children, 758 children were admitted with acute malaria to Kilifi County Referral and Teaching Hospital (KCRTH), while 208 children presented with mild malaria at outpatient clinics within Kilifi County (**Figure 1A**).

Parasite *var* gene expression profiles were generated using either DBLα-tag sequencing (Bull et al., 2005) or RT-qPCR (Lavstsen et al., 2012; Abdi et al., 2016). DBLα-tag sequencing data were available for 898 samples, including 716 hospital admissions and 182 from outpatient cases, while RT-qPCR *var* expression profile were available for 768 samples, including 561 hospital admissions and 207 outpatient cases (**Figure 1A**). The two datasets substantially overlap, with 702 samples having both DBLα-tag and RT-qPCR data (**Figure 1A**).

Within the DBLα-tag dataset, 216 children were admitted with moderate malaria and 500 with severe malaria (**Figure 1B**). Among the children with severe malaria, 249 presented with impaired consciousness, 41 with respiratory distress, 76 with severe malarial anaemia and 134 with more than one syndrome (overlapping syndrome group) (**Figure 1B**).

In the RT-qPCR dataset, 152 children were admitted with moderate malaria and 409 with severe malaria (**Figure 1C**). Among these severe cases, 206 had impaired consciousness, 29 had respiratory distress, 59 had severe malarial anaemia and 115 had overlapping severe malaria syndromes (**Figure 1C**). For subsequent analysis, children with mild or moderate malaria were grouped as having non-severe malaria (NSM). Children with SMA were the youngest (P<0.001), whereas those with RD had the highest median peripheral blood parasitaemia (P=0.037) (**Table S1A&B).**

The mean age of children increased across transmission periods, from 33.2 months in the pre-decline period to 43.7 months during the decline period and 54.4 months in the post-decline period (**Figure S1)**. We therefore compared *var* gene expression across transmission periods using both unadjusted and age-adjusted linear regression models. No consistent differences in the expression of the *var* gene subsets were observed across transmission periods, except for group B and C that showed increasing trends with declining transmission (**Table S2).**

To maximise the use of the available data, the DBLα-tag and RT-qPCR datasets were analysed and the results presented independently.

**Figure 1:**
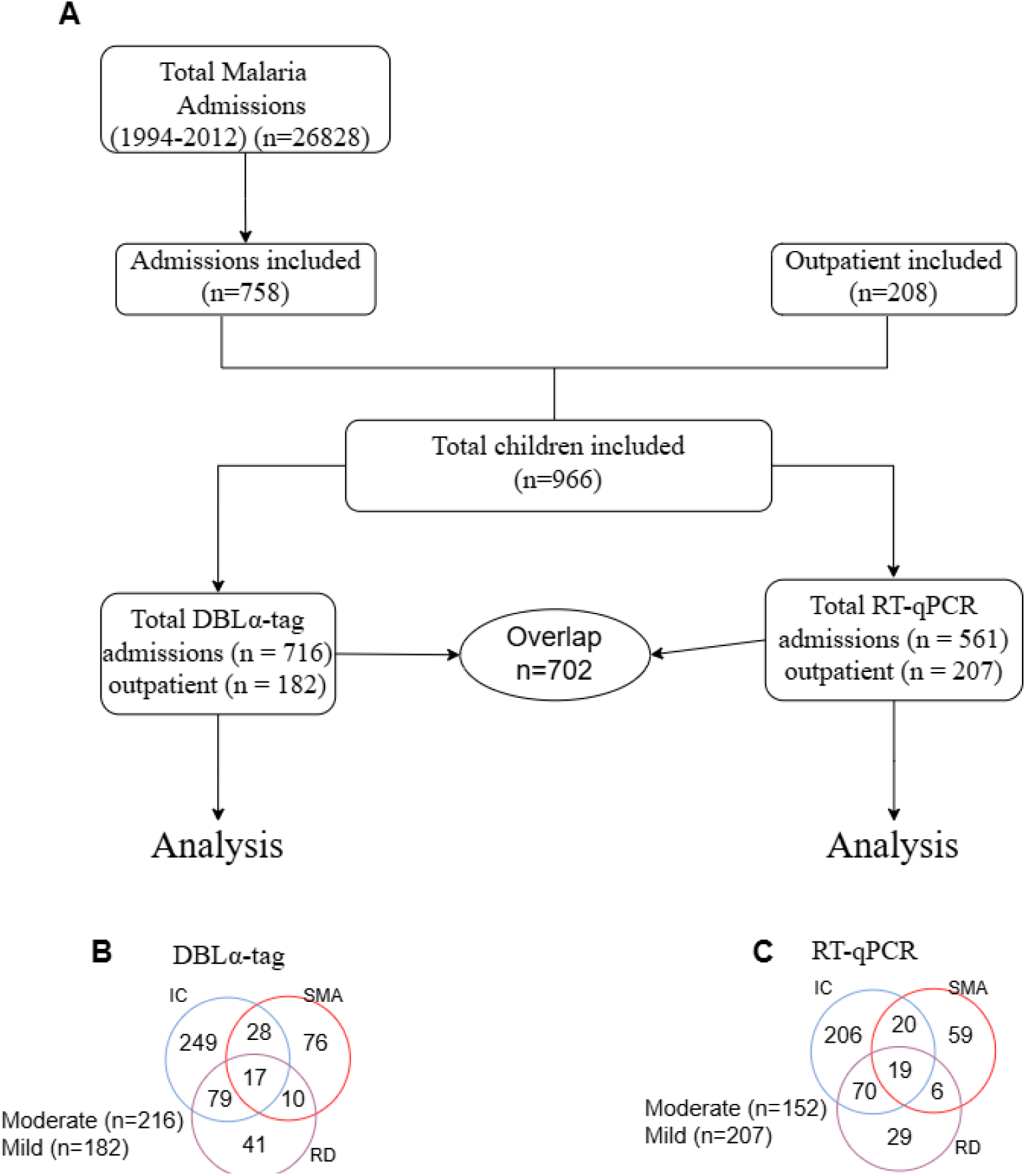
Sample inclusion and syndrome classification. **A)**. Flowchart showing the children included in the study. Children were included based on the availability of DBLα-tag and/or RT-qPCR data, together with complete participant information required to define the clinical categories, age and peripheral blood parasitaemia**. (B-C)** Venn diagrams showing the distribution of children presenting with single and overlapping severe malaria syndromes among those with **(B)** DBLα-tag and **(C)** RT-qPCR data. IC: impaired consciousness, RD: Respiratory distress, SMA: severe malarial anaemia. Mild and moderate are combined to form non-severe malaria group (NSM)

### Impaired consciousness was associated with cys2 and group A-like *var* gene expression

We used DBLα-tag sequencing to capture the breadth and relative proportions of expressed *var* repertoires. Sequence tags were classified according to cysteine residue number and PoLV motifs into cys2, cys4, cysX, and cp1–cp6 categories, with bs1_cys2 defined as “group A-like” (**Table 1**) (Bull et al., 2005). Because cys2 and cys4 proportions are inversely correlated, the analysis focused on cys2 and its constituent subgroups.

**Table 1:** DBLα-tags sequence groupings and common names used in the text.

| Grouping | Original name | Common name | No. of cysteine residues | Unique motif /feature |
| --- | --- | --- | --- | --- |
| cp | Group 1 (cys2/ MFK+&REY-) | cp1 | 2 | MFK |
|  | Group 2 (cys2/ MFK-&REY+) | cp2 | 2 | REY |
|  | Group 3 (cys2/ MFK-&REY-) | cp3 | 2 | None |
|  | Group 4 (cys4/ MFK-&REY-) | cp4 | 4 | None |
|  | Group 5 (cys4/ MFK-&REY+) | cp5 | 4 | REY |
|  | Group 6 (cysX) | cp6 | 1,3,X | Variable |
| bs | bs1_Cys2 | group A-like | 2 | bs1 |

cp: cysteine/position of limited variation, bs: block sharing, X: any number other than 2 or 4. All cys4 DBLα-tags are either group B or C while most of cys2 are group A *var* genes but not all group A are cys2

We fitted five independent multiple linear regression models predicting the expression of cys2 *var* genes and its subtypes (bs1_cys2, cys2 MFK+REY-, cys2 MFK-REY+, cys2 MFK-REY-) hereafter referred to as group A-like, cp1, cp2 and cp3 respectively (**Table 1**) as dependent variables. The explanatory variables included the three primary clinical syndromes (impaired consciousness [IC], respiratory distress [RD], and severe malarial anaemia [SMA]), along with age and peripheral blood parasite density.

Compared with children without IC (including the non-severe cases), IC was significantly and positively associated with cys2 [beta (β) (95%CI): 0.09 (0.05, 0.13), P_adjusted_= 0.0001], group A-like [β(95%CI): 0.08(0.04, 0.11), P_adjusted_=0.0001], and cp1 [β(95%CI): 0.08(0.05, 0.11), P_adjusted_= 0.000001] (**Figure 2A, Table S3**). RD did not show significant association with the expression of any of the DBLα-tag-defined *var* subtypes, however, compared with the other syndromes, RD showed stronger association with cp3 (**Figure 2B**, **Table S3**). SMA showed association with group A-like expression, however, this association was no long significant after multiple testing correction [β(95%CI): 0.06 (0.01, 0.12), P=0.04]. (**Figure 2C**, **Table S3**).

To determine whether these associations were driven by specific syndromes rather than overlapping presentations, we repeated the above analysis after excluding children with overlapping severe syndromes. After multiple testing correction, cys2, group A-like and cp1 remained significantly associated with IC (P_adjusted_= 0.0004, 0.0002 and 0.0002, respectively) (**Figure 2D, Table S3)**. Consistent with the primary analysis, RD was not associated with the expression of any DBLα-tag-defined *var* subtype (**Figure 2E, Table S3**), whereas SMA showed an unadjusted association with increased group A-like expression (P = 0.03), which did not remain significant after multiple testing correction (**Figure 2F**, **Table S3**).

Because 182 of the 898 DBLα-tag expression profiles included in this study were also analysed in a previous publication (Warimwe et al., 2009), we performed a sensitivity analysis after excluding these overlapping samples. The restricted dataset comprised 716 children, including 304 with NSM, 207 with IC, 32 with RD, 67 with SMA and 106 with overlapping syndromes. Reanalysis of this independent dataset yielded findings consistent with the primary analysis, with the association between RD and cp3 reaching significance (**Figure S2**), indicating that the observed associations were not driven by the subset of previously analysed samples.

**Figure 2.**
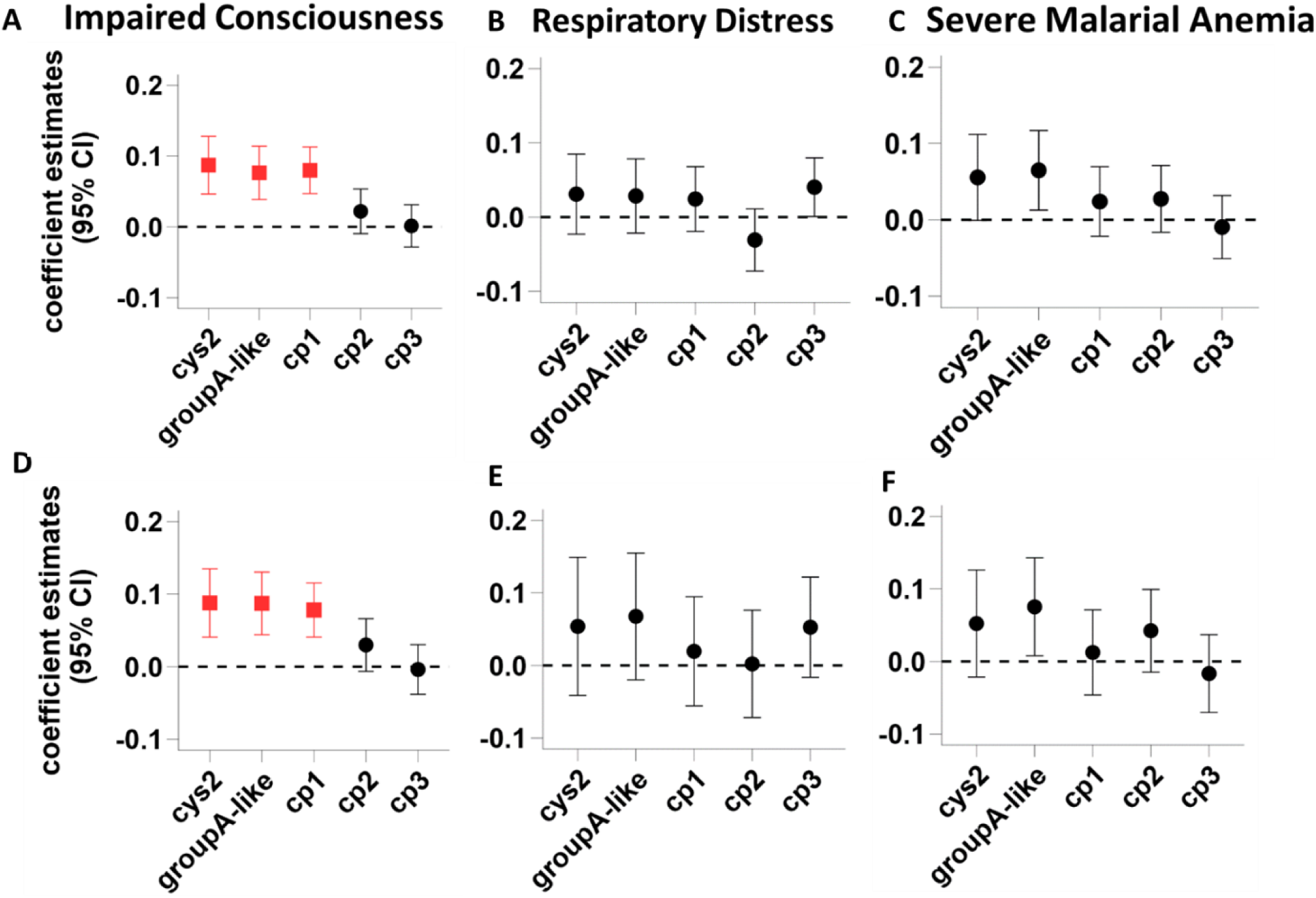
Associations between *var* gene expression and severe malaria syndromes using DBLα-tag sequencing. **A-F**). Multiple linear regression estimates for associations between *var* variants (cys2, group A-like, cp1, cp2, and cp3) and severe malaria syndromes; impaired consciousness (IC), respiratory distress (RD), and severe malarial anaemia (SMA). Five independent models were fitted with each *DBLα-tag group* as outcome and IC, RD, and SMA as explanatory variables, adjusting for age and peripheral blood parasite density. **A-C).** Analyses including all *DBLα-tag sequenced* samples (n = 898). **D-F).** Analyses restricted to samples with a single severe malaria syndrome (n = 764). Within each row, panels show association with IC (left), RD (middle), and SMA (left). Horizontal error bars represent 95% confidence intervals and dashed horizontal line denotes the null effect (β=0). Red squares indicate significant positive associations, and black circles indicate associations that are not significant after multiple testing correction (FDR<0.05), IC (n=249), RD (n=41), SMA (n=76) overlapping syndromes (n=134) moderate (n=216), and mild (n=182)

### IC showed the most robust association with group A and DC8 transcript levels

To complement the DBLα-tag sequencing analysis, we analysed RT-qPCR data targeting specific *var* gene subgroups, including group A (gpA1, gpA2, dc13), DC8 (dc8-1 to dc8-4), DC9 (dc9), and broadly group B (gpB) and C (gpC) transcripts (**Table S4).** For each transcript, we fitted multiple linear regression model with transcript abundance as the outcome and IC, RD and SMA included simultaneously as explanatory variables while adjusting for age and peripheral parasitaemia.

In the primary analysis, which includes overlapping syndromes, IC remained significantly associated after multiple testing correction with higher transcript levels measured using gpA1 [β (95%CI): 0.38 (0.12, 0.63), P_adjusted_=0.042], gpA2 [β (95%CI): 0.33 (0.07, 0.59), P_adjusted_=0.04] and dc8-3 [β (95%CI): 0.40 (0.10, 0.70), P_adjusted_=0.04] primers (**Figure 3A**, **Table S5**), as well as with higher median group A expression (gpA-median) [β (95%CI): 0.33(0.07, 0.58), P_adjusted_=0.037]. IC also showed significant association with the median DC8 expression (dc8-median) in unadjusted analysis but did not remain significant after multiple testing correction (**Figure 3A**, **Table S5**). Consistent with DBLα-tag sequencing analysis, RD was not associated with the expression of any targeted *var* transcript (**Figure 3B**). Conversely, SMA was negatively associated with dc9 [β (95%CI): -0.50(-0.89, -0.12), P=0.01] and gpB expression, with the latter remaining significant after multiple testing correction [β (95%CI): -0.50(-0.82, -0.18), P_adjusted_=0.03]. (**Figure 3C, Table S5**).

To determine whether these associations were influenced by the overlapping clinical syndromes, we repeated the analysis after excluding children with multiple severe syndromes. IC remained significantly associated with gpA1 [β (95%CI) 0.52 (0.22, 0.81), P_adjusted_=0.002], gpA2 [β (95%CI) 0.55 (0.25, 0.84), P_adjusted_=0.002], gpA-median [β (95%CI) 0.50 (0.21, 0.79), P_adjusted_=0.002] and dc8-3 [β (95%CI) 0.68 (0.34, 1.02), P_adjusted_=0.001]. Furthermore, IC became significantly associated with dc8-2 [β (95%CI) 0.62 (0.26, 0.97), P_adjusted_=0.002] and dc8-median [β (95%CI) 0.53 (0.21, 0.84), P_adjusted_=0.002] while the association with dc8-4 was no longer significant after multiple test correction (**Figure 3D, Table S5**).

For RD, significant positive associations were observed with gpA2 [β (95%CI): 0.66(0.11, 1.2), P_adjusted_ =0.019], and gpA-median [β (95%CI): 0.56(0.01, 1.1), P_adjusted_ =0.046], while associations with gpA1, dc8-2, and dc8-3 were significant before multiple testing correction but after adjusting significance was lost (**Figure 3E, Table S5**).

SMA remained positively associated with gpA2 [β (95%CI): 0.77 (0.29, 1.24), P_adjusted_=0.02], gpA-median [β (95%CI): 0.63 (0.15, 1.11), P_adjusted_=0.04], dc8-1 [β (95%CI): 0.75 (0.15, 1.35), P_adjusted_=0.04], dc8-3 [β (95%CI): 0.71 (0.17, 1.26), P_adjusted_=0.04]. Although gpA1, dc8-2 and dc8-median showed unadjusted association, these did not remain significant after multiple testing correction. Conversely, the negative associations with gpB and dc9 observed in the primary analysis were no longer significant after excluding children with overlapping syndromes (**Figure 3F, Table S5**).

**Figure 3.**
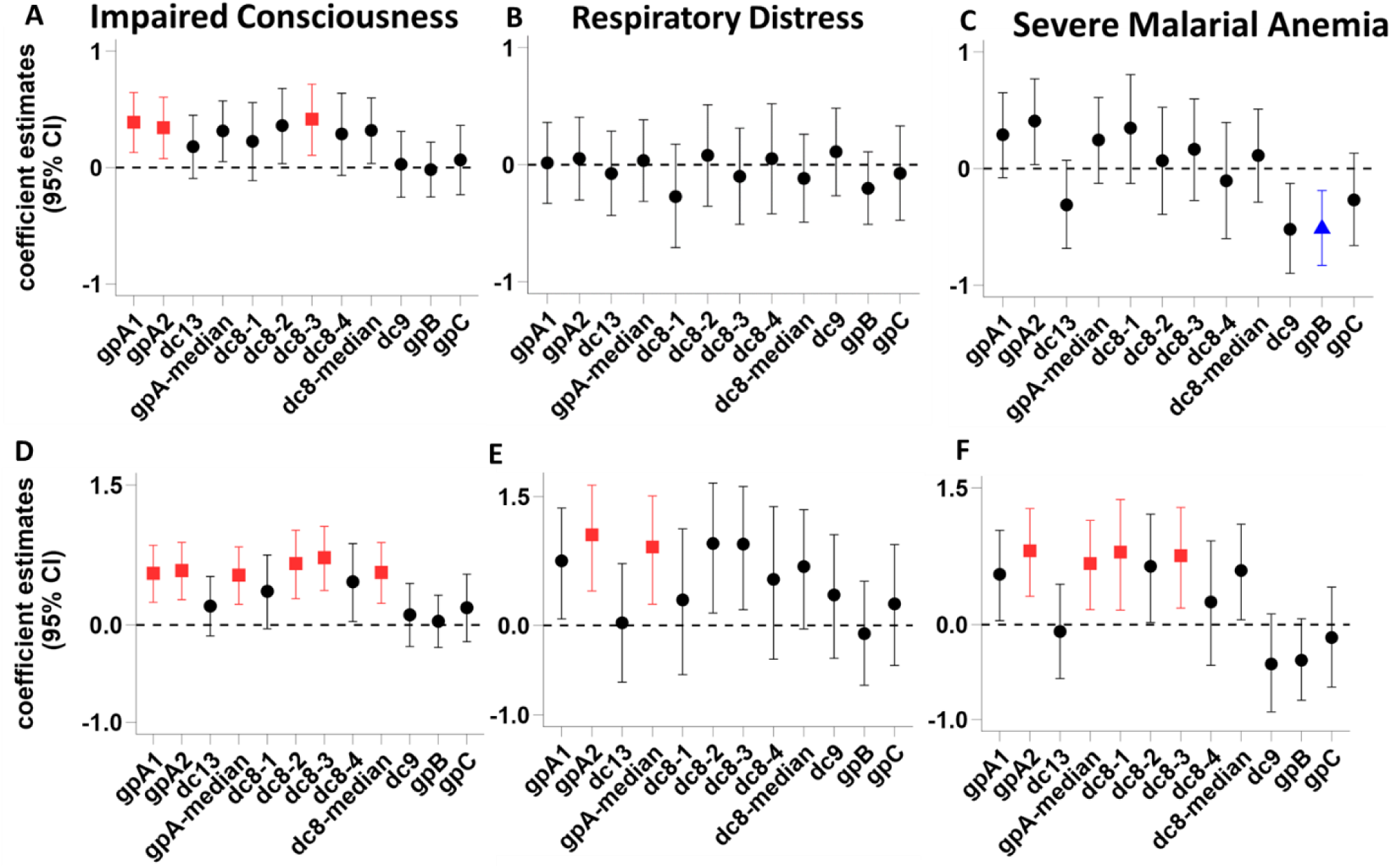
Associations between *var* gene expression measured by RT-qPCR and severe malaria syndromes. **A-F**). Multiple linear regression estimates for associations between *var* variants (gpA1-gpC) and severe malaria syndromes; Impaired Consciousness (IC), Respiratory Distress (RD), and Severe Malarial Anaemia (SMA). Thirteen independent models were fitted with each *var* transcript measure as outcome and IC, RD, and SMA as explanatory variables, adjusting for age and peripheral blood parasite density. **A-C).** Analyses including all RT-qPCR samples (n = 768). **D-F).** Analyses restricted to samples with a single severe malaria syndrome (n = 653). Within each row, panels show association with IC (left), RD (middle), and SMA (left). Horizontal error bars represent 95% confidence intervals and dashed horizontal line denotes the null effect (β=0). Red squares indicate significant positive associations, blue triangles indicate significant negative association, and black circles indicate non-significant association after multiple testing correction (FDR<0.05). IC (n=206), RD (n=29), SMA (n=59) overlapping syndromes (n=115) moderate (n=152), and mild (n=207)

### DBLα-tag-based prediction recapitulates the association of IC with group A, EPCR- and EPCR-ICAM-1 binding *var* gene expression

To determine whether sequence-based prediction from DBLα-tag recapitulated the *var* expression patterns observed by direct transcript quantification, we predicted upstream sequence (UPS) groups and *Pf*EMP1 domain structures from DBLα-tag sequences using recently developed bioinformatics tools. cUPS (Tan et al., 2025) and UpsML (Pangilinan et al., 2026) were used to predict UPS groups (UPSA, UPSB, and UPSC), while Varia (Mackenzie et al., 2022) was used to infer *Pf*EMP1 domain structures. Predicted domain structures were subsequently classified into domain cassettes (DCs) **(Table S6A)** (Rask et al., 2010) and binding phenotypes (**Table S6B**) (Gamain et al., 2002; Robinson et al., 2003; Turner et al., 2013; Smith, 2014; Jespersen et al., 2016) The proportion of DBLα-tag sequences assigned to each UPS group, domain cassette, or receptor binding phenotype (EPCR, dual EPCR-ICAM-1, PCAM, gC1qR, CD36 and CSA) was calculated and used as the outcome variable in multiple linear regression models, with IC, RD, and SMA as explanatory variables, adjusting for age and peripheral parasite density.

Consistent with the RT-qPCR findings, IC was significantly associated with a higher predicted UPSA proportion and lower UPSB proportion using both cUPS and UPSML (**Figure 4A-B, top panel**). For RD, UPSML predicted a significant lower UPSB proportion after multiple-testing correction (**Figure 4A-B**, middle). In contrast, SMA was associated with a lower predicted UPSB proportion using cUPS only, with no significant association detected for the other predicted UPS groups (**Figure 4A, bottom panel**).

At the predicted binding-phenotype level, IC was associated with higher proportional expression of variants associated with EPCR-binding [CIDRα1.1/1.4-1.8, β (95% CI): 0.06 (0.02, 0.10), P_adjusted_ = 0.01] and dual EPCR-ICAM-1 binding [DBLa1.4/7-CIDRa1.4/6-DBLb3, β (95% CI): 0.06 (0.02, 0.10), P_adjusted_ = 0.01]. Conversely, IC was associated with a lower proportion of predicted CD36-binding variants [DBLa0-CIDRa2-9, β (95% CI) −0.05 (−0.10, −0.01), P_adjusted_ = 0.04) (**Figure 4C top panel**). No predicted binding phenotype was associated with RD (**Figure, 4C middle**), and SMA (**Figure 4C bottom).** Analysis of individual predicted domain cassettes showed no significant association with IC, RD and SMA (**Figure S3**).

Finally, sequence-based prediction showed concordance with *var* subtypes quantified by RT-qPCR (**Figure 4D**). In particular, predicted UPSA and UPSB proportions correlated with group A and group B transcript quantity. Similarly, the predicted EPCR or dual EPCR-ICAM-1 binders showed positive association with group A and DC8 transcript levels, supporting the ability of DBLα-tag based approach to recover major features of the expressed *Pf*EMP1 repertoire.

Overall, IC showed the most consistent and robust association with increased group A and DC8 transcript expression as well as with higher predicted expression of variants containing domains associated with EPCR and dual EPCR-ICAM-1 binding phenotypes. In contrast, associations with SMA emerged only after excluding children with overlapping severe malaria syndromes and remained less consistent, whereas RD showed little evidence of consistent association with expression of the targeted *var* transcripts.

**Figure 4.**
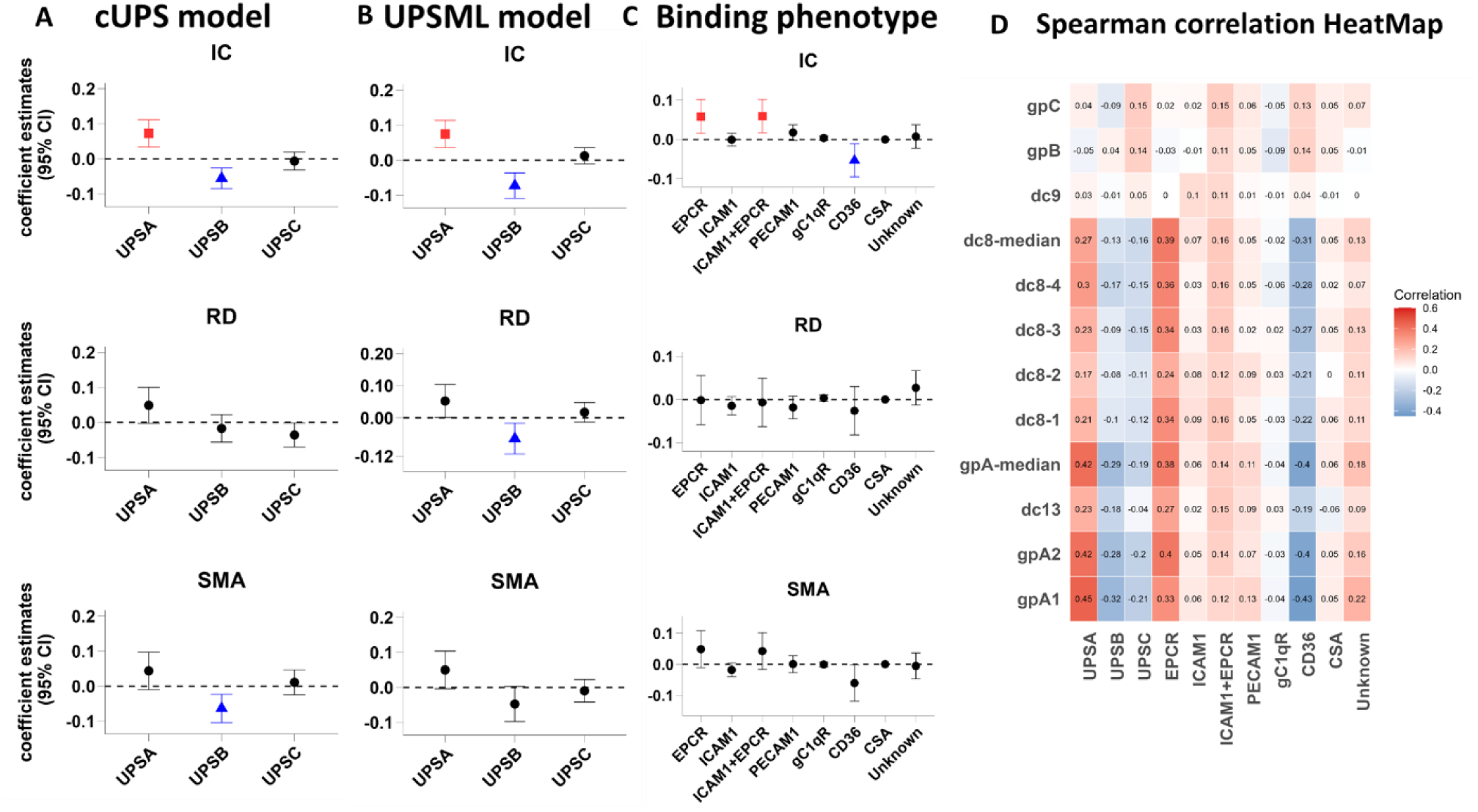
DBLα-tag based prediction of UPS groups, binding phenotypes in relation to disease severity. **A-C**). Multiple linear regression estimates for associations between IC (top), RD (middle), and SMA (bottom) and the proportion of predicted UPS groups or *Pf*EMP1 binding phenotypes. UPS groups were predicted using the cUPS algorithm **(A)** and UPSML **(B),** while *Pf*EMP1 binding phenotypes were inferred from predicted domain architecture. Red squares indicate significant positive associations, blue triangles indicate significant negative associations, and black circles indicate non-significant association after multiple testing correction (FDR<0.05). Horizontal error bars represent 95% confidence intervals and dashed horizontal line denotes the null effect (β=0). Models were adjusted for age and peripheral parasite density. **D).** Spearman correlation heatmap comparing RT-qPCR-derived transcript levels (rows) with DBLα-tag-predicted UPS groups and binding phenotypes (columns). Cell values represent the spearman correlation coefficients and colour scale indicates Spearman’s rho, with red denoting positive and blue denoting negative correlation.

### SMA parasites have a higher homogenous *var* expression profile among admissions

To determine whether parasites associated with different severe malaria syndromes differ in the number of distinct *var* genes they express, we compared *var* expression homogeneity (VEH) across clinical malaria presentations. Higher VEH values indicate that a small number of *var* genes dominate the expressed repertoire in a sample, whereas low VEH reflects a more diverse *var* expression profile. VEH was calculated from the expressed sequence tag data as previously described (Warimwe et al., 2013). In 752 children presenting with single syndromes, parasites from children with SMA exhibited the highest VEH, with significantly higher values than those observed in IC (P = 0.001), moderate (P= 0.001), and mild (P= 0.02) malaria cases (**Figure 5A**). Consistent with this, children with SMA showed higher VEH values than non-SMA cases (P = 0.001) (**Figure 5B**).

To explore factors underlying the elevated VEH observed in SMA, we examined associations with parasite and host variables. VEH was negatively correlated with examined *var* subtypes quantified by RT-qPCR. Notably, group A *var* subtypes showed the weakest negative correlations, whereas group B and C showed the strongest negative correlation with VEH (**Figure 5C**). *Pf*Sir2a, which was associated with dysregulation of *var* gene expression, particularly group B *var* genes (Merrick et al., 2012; Abdi et al., 2016, 2023), showed a weak non-significant negative correlation with VEH (**Figure 5C**). However, *Pf*Sir2a expression was significantly lower in SMA compared with IC (*p* = 0.03) (**Table S7**).

We next examined whether antibody recognition of IEs was associated with VEH. For each patient, we measured anti-infected erythrocyte (α-IE) IgG levels against trophozoite-infected erythrocytes from two heterologous isolates at the time of sampling using flow cytometry. The median α-IE level was positively associated with VEH (rho = 0.14, p = 0.001) (**Figure 5C**), consistent with previous work suggesting that antibody-mediated selection can favour more homogeneous *var* expression profiles (Warimwe et al., 2013). However, children with SMA were younger (**Table S1**) and had the lowest α-IE levels (**Figure 5D**). Reported fever duration prior to admission was longest in SMA and shortest in IC (**Figure 5E**), consistent with SMA reflecting a more prolonged infection, although reported fever duration itself was not associated with VEH (rho = 0.004, *p* = 0.94) (**Figure 5C**). In contrast, febrile temperature showed weak but significant negative association with VEH (rho = −0.09, *p* = 0.03) (**Figure 5C**) and was lowest in SMA (median [IQR] 38°C [37.2-38.7]; **Table S8**), consistent with reduced *Pf*Sir2a expression in this group (Abdi et al., 2023).

**Figure 5:**
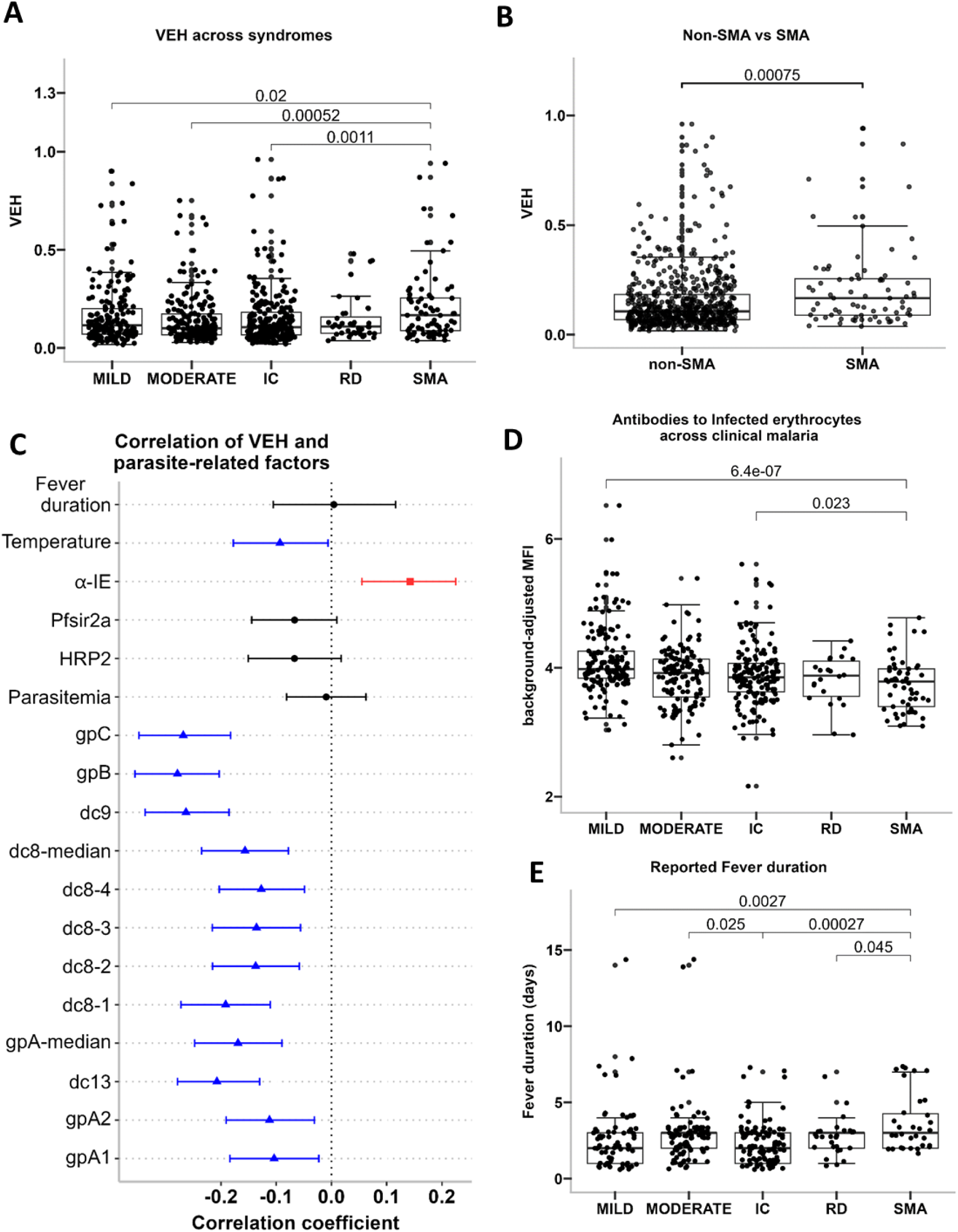
*Var* gene expression homogeneity (VEH) in relation to clinical malaria, parasite and host factors. **A).** Distribution of *Var* gene Expression Homogeneity (VEH) values across pure clinical malaria groups in children. The y-axis shows VEH scores (higher values show more homogenous *var* expression); x-axis show clinical presentation. **B)**. Comparison of VEH between SMA (n=59) and non-SMA cases comprising all other severe and non-severe cases (n=532). ***C)***. Spearman’s correlation (95% bootstrap confidence intervals) between VEH and parasite- and host-related factors. Red squares indicate significant positive correlations, blue triangles indicate significant correlations and black circles indicate non-significant correlations. **D).** Background-adjusted MFI of antibody recognition of infected erythrocytes (α-IE) across clinical malaria groups. **E)**. Caregiver-reported fever duration (days) prior to admission. In **A, B, D**, and **E,** individual observations are overlaid on boxplots; horizontal brackets indicate statistically significant pairwise comparisons (Wilcoxon rank-sum test), with corresponding p-values shown. IC, impaired consciousness; RD, respiratory distress; SMA, severe malarial anaemia

### Group A *var* genes and rosetting are differentially associated with severe malaria syndromes

Rosette frequency was assessed in a subset of children for whom rosette counts were available from *ex vivo* assays of parasite isolates (n = 273). For each isolate, the proportion of infected erythrocytes forming rosettes was quantified and used as a continuous measure of rosetting. Rosette frequency was significantly higher among children with severe malaria than among those with non-severe malaria [median (IQR): 7.0% (1.0-21.0) vs 3.7% (0-13.0), Wilcoxon rank-sum test, p = 0.013] **(Figure S4)**.

To evaluate the relationship between rosette frequency and specific severe malaria syndrome, pairwise Wilcoxon rank-sum test was performed comparing rosette frequency between children with and without each syndrome. Rosette frequency was significantly higher in children with RD than in those without RD (p = 0.002), and in children with SMA than in those without SMA (p = 0.04) but did not differ significantly between IC and non-IC cases (p = 0.10) (**figure 6A**).

To account for overlapping clinical presentations while adjusting for age and peripheral parasite density, a linear regression analysis was fitted with rosette frequency as the outcome, and IC, RD, and SMA as predictors. Rosette frequency remained positively associated with RD [β(95%CI) 0.11 (0.01, 0.20), P=0.025] and SMA [β(95%CI) 0.12 (0.03, 0.23), P=0.015], but not with IC [β(95%CI) 0.03 (-0.04, 0.10), P=0.40] (**Figure 6B**).

We further show that rosette frequency was positively correlated with the expression of group A, DC8 *var* variants, as well as predicted rosetting-associated domain cassettes (8,11 and 15), UPSA, and binding phenotypes [EPCR, dual EPCR-ICAM-1 and PECAM], while gpB transcript levels, and predicted UPSB, UPSC, CD36 binding phenotype were negatively correlated with rosette frequency (**Figure 6C**).

To determine whether RD and SMA associations with rosetting were independent of *var* gene expression, we fitted logistic regression models for each syndrome with rosette frequency as a predictor while adjusting separately for expression levels of *var* gene subtypes that showed positive associations with RD or SMA in the primary analysis (group A-like, gpa1,gpA2,gpA-median, dc8-1,dc8-2,dc8-3, dc8-4 and dc8-median), as well as predicted rosetting-associated domain cassettes (DC8,DC11,DC15 and DC16) (McLean et al., 2025).

For RD, the association with rosetting remained significant after adjustment for all examined *var* gene subsets and predicted rosetting-associated domain cassettes (**Figure 6D**; **Table 2**). Adjusted odds ratios ranged from 3.14 to 4.56. Similarly, rosetting remained significantly associated with SMA after adjustment for group A-like, DC8-related subsets, and predicted rosetting-associated domain cassettes (DC8, DC11, DC15, and DC16) (**Figure 6E**; **Table 2**). However, the association was attenuated and no longer statistically significant after adjustment for gpA1, gpA2, or gpA-median expression. These findings suggest that the relationship between rosetting and SMA may be partially mediated through specific group A var gene subsets, whereas the association between rosetting and RD appears to be largely independent of the *var* gene variants examined.

**Figure 6:**
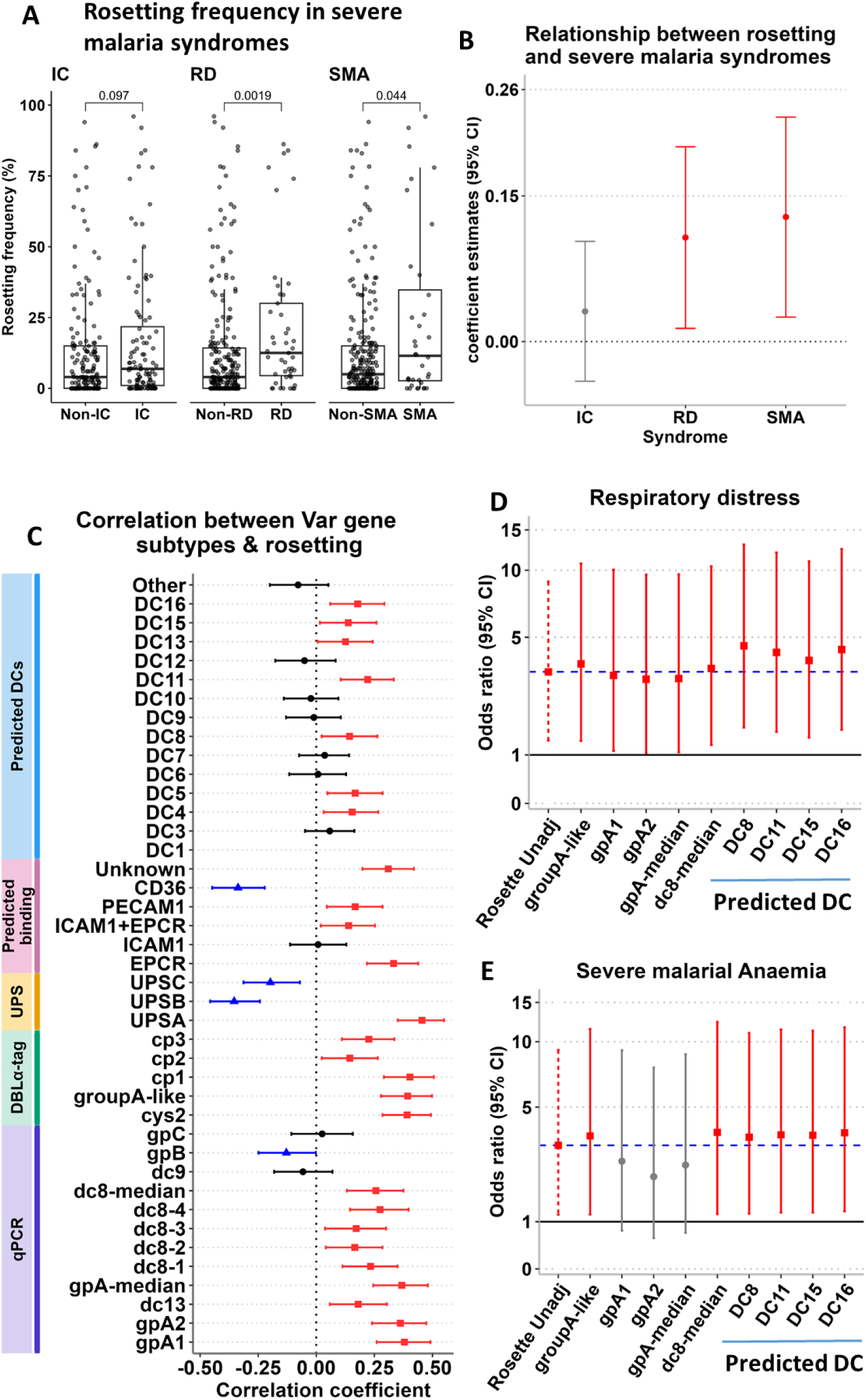
Relationship between rosetting, *var* gene expression and severe malaria. **A)**. Rosetting frequency among children with and without specific severe malaria syndromes: impaired consciousness (IC), respiratory distress (RD), and severe malarial anaemia (SMA). Boxplots show the distribution of rosetting frequencies, with individual observations overlaid. p values were calculated using the Wilcoxon rank-sum test. IC (n = 71), non-IC (n = 202); RD (n = 17), non-RD (n = 256); SMA (n = 21), and non-SMA (n = 252).**B).** Regression coefficients (β) and 95% confidence intervals (CI) for the association between rosette frequency and severe malaria syndromes. **C)**. Spearman correlation coefficients (95% bootstrap confidence intervals) between rosette frequency and expression levels of different *var* gene subgroups measured or predicted using complementary approaches: RT-qPCR (qPCR), DBLα-tag sequencing, predicted upstream promoter sequence (UPS) classes, predicted receptor-binding phenotypes, and predicted domain cassettes (DCs). Red squares indicate significant positive correlations, blue triangles indicate significant negative correlations, and black circles indicate non-significant correlations. **D-E).** Odds ratios (95% CI) for associations between rosette frequency and **(D)** Respiratory distress and **(E)** Severe malarial anaemia, before and after adjustment for individual rosetting-associated *var* gene features. Each model included rosette frequency and one *var* gene feature at a time. The blue dashed line indicates the unadjusted odds ratio for rosetting.

**Table 2:** Relationship between rosetting and *var* gene subtypes that showed positive association with RD or SMA in the primary analysis (group A-like, gpa1,gpA2,gpA-median, dc8-1,dc8-2,dc8-3, dc8-4 and dc8-median) as well as predicted rosetting-associated Domain cassettes (8,11,15 and 16) in different malaria syndromes using logistic regression models. Models 1-14 are unadjusted while models 15 to 27 are adjusted for *var* gene expression levels. Bold p values are significant at alpha less than 0.05.

| Model | Variables | A: Impaired consciousness |  | B: Respiratory distress |  | C: Severe malaria anaemia |  |
| --- | --- | --- | --- | --- | --- | --- | --- |
|  |  | OR (95% CI) | p value | OR (95% CI) | p value | OR (95% CI) | p value |
| **1**<br>**A,B,C** | Rosetting | 2.03 (0.87, 4.79) | 0.10 | 4.39 (1.6, 12.03) | **<0.001** | 3.56 (1.17, 10.71) | **0.02** |
| **2**<br>**A,B,C** | Group A-like | 5.44 (2.23, 13.8) | **<0.001** | 2.69 (0.89, 8.25) | 0.80 | 1.44 (0.42, 4.87) | 0.56 |
| **3**<br>**A,B,C** | gpA1 | 1.36 (1.15, 1.64) | **<0.001** | 1.16 (0.93, 1.47) | 0.19 | 1.42 (1.08, 1.92) | **0.02** |
| **4**<br>**A,B,C** | gpA2 | 1.24 (1.05, 1.48) | **0.01** | 1.2 (0.96, 1.52) | 0.12 | 1.71 (1.27, 2.39) | **<0.001** |
| **5**<br>**A,B,C** | gpA-median | 1.29 (1.09, 1.55) | **<0.001** | 1.19 (0.96, 1.51) | 0.121 | 1.5 (1.13, 2.04) | **0.01** |
| **6**<br>**A,B,C** | dc8-1 | 1.14 (1.01, 1.3) | **0.04** | 1 (0.85, 1.18) | 0.98 | 1.06 (0.88, 1.28) | 0.56 |
| **7**<br>**A,B,C** | dc8-2 | 1.17 (1.04, 1.33) | **0.01** | 1.08 (0.92, 1.28) | 0.33 | 1.1 (0.92, 1.33) | 0.29 |
| **8**<br>**A,B,C** | dc8-3 | 1.2 (1.05, 1.39) | **0.01** | 0.98 (0.83, 1.17) | 0.86 | 1.09 (0.89, 1.35) | 0.41 |
| **9**<br>**A,B,C** | dc8-4 | 1.17 (1.03, 1.34) | **0.02** | 1.18 (0.99, 1.4) | 0.07 | 1.1 (0.9, 1.34) | 0.36 |
| **10**<br>**A,B,C** | dc8-median | 1.28 (1.09, 1.5) | **<0.001** | 1.08 (0.89, 1.32) | 0.46 | .09 (0.87, 1.39) | 0.45 |
| **11**<br>**A,B,C** | DC8 | 0.96 (0.27, 3.35) | 0.95 | 0.27 (0.04, 1.51) | 0.16 | 2.11 (0.35, 11.59) | 0.40 |
| **12**<br>**A,B,C** | DC11 | 0.58 (0.1, 3.25) | 0.54 | 0.87 (0.08, 7.58) | 0.90 | 1.07 (0.07, 11.57) | 0.96 |
| **13**<br>**A,B,C** | DC15 | 1.12 (0.21, 5.59) | 0.89 | 7.76 (1.22, 48.3) | **0.03** | 0.98 (0.06, 8.77) | 0.99 |
| **14**<br>**A,B,C** | DC16 | 1.04 (0.22, 4.58) | 0.96 | 0.52 (0.04, 3.63) | 0.56 | 0.71 (0.05, 5.5) | 0.77 |
| **15**<br>**A,B,C** | Rosetting | 1.19 (0.46, 2.98) | 0.71 | 3.74 (1.28, 10.75) | **0.01** | 3.61 (1.12, 11.46) | **0.03** |
|  | Group A-like | 4.7 (1.85, 12.37) | **<0.001** | 1.89 (0.58, 6.09) | 0.29 | 1.02 (0.27, 3.7) | 0.97 |
| **16**<br>**A,B,C** | Rosetting | 1.34 (0.51, 3.5) | 0.55 | 3.28 (1.05, 10.09) | **0.04** | 2.66 (0.75, 9.2) | 0.12 |
|  | gpA1 | 1.33 (1.11, 1.61) | **<0.001** | 1.08 (0.85, 1.38) | **0.54** | 1.34 (1, 1.84) | 0.06 |
| **17**<br>**A,B,C** | Rosetting | 1.57 (0.6, 4.06) | 0.35 | 3.14 (1, 9.61) | **0.04** | 2.17 (0.59, 7.69) | 0.23 |
|  | gpA2 | 1.2 (1, 1.44) | 0.05 | 1.12 (0.89, 1.44) | 0.35 | 1.64 (1.2, 2.31) | **<0.001** |
| **18**<br>**A,B,C** | Rosetting | 1.48 (0.57, 3.84) | 0.42 | 3.16 (1.01, 9.64) | **0.04** | 2.53 (0.71, 8.83) | 0.15 |
|  | gpA-median | 1.25 (1.05, 1.51) | **0.01** | 1.12 (0.89, 1.43) | 0.35 | 1.42 (1.06, 1.95) | **0.02** |
| **19**<br>**A,B,C** | Rosetting | 1.81 (0.72, 4.57) | 0.21 | 3.75 (1.25, 11.11) | **0.02** | 3.78 (1.13, 12.44) | **0.03** |
|  | dc8-1 | 1.12 (0.98, 1.28) | 0.09 | 0.97 (0.82, 1.15) | 0.70 | 1.03 (0.85, 1.25) | 0.76 |
| **20**<br>**A,B,C** | Rosetting | 1.87 (0.74, 4.71) | 0.18 | 3.56 (1.19, 10.45) | **0.02** | 3.79 (1.14, 12.38) | **0.03** |
|  | dc8-2 | 1.16 (1.02, 1.32) | **0.02** | 1.07 (0.91, 1.27) | 0.41 | 1.09 (0.91, 1.32) | 0.34 |
| **21**<br>**A,B,C** | Rosetting | 1.86 (0.74, 4.7) | 0.19 | 3.7 (1.24, 10.85) | **0.02** | 3.8 (1.14, 12.48) | **0.03** |
|  | dc8-3 | 1.19 (1.04, 1.38) | **0.02** | 0.97 (0.81, 1.16) | 0.70 | 1.08 (0.88, 1.35) | 0.47 |
| **22**<br>**A,B,C** | Rosetting | 1.68 (0.66, 4.27) | 0.27 | 3.2 (1.04, 9.55) | **0.04** | 3.67 (1.1, 12.07) | **0.03** |
|  | dc8-4 | 1.16 (1.02, 1.34) | **0.03** | 1.15 (0.96, 1.37) | **0.13** | 1.07 (0.88, 1.32) | 0.50 |
| **23**<br>**A,B,C** | Rosetting | 1.71 (0.67, 4.35) | 0.26 | 3.55 (1.18, 10.45) | **0.02** | 3.76 (1.13, 12.33) | **0.03** |
|  | dc8-median | 1.26 (1.07, 1.48) | **0.01** | 1.05 (0.86, 1.29) | 0.64 | 1.07 (0.85, 1.36) | 0.56 |
| **24**<br>**A,B,C** | Rosetting | 1.7 (0.71, 4.1) | 0.23 | 4.56 (1.6, 13) | **<0.001** | 3.56 (1.14, 11) | **0.03** |
|  | DC8 | 0.91 (0.25, 3.21) | 0.88 | 0.2 (0.03, 1.22) | 0.10 | 1.99 (0.32, 11.26) | 0.44 |
| **25**<br>**A,B,C** | Rosetting | 1.77 (0.73, 4.31) | 0.20 | 4.25 (1.5, 12) | **0.01** | 3.66 (1.17, 11.4) | **0.02** |
|  | DC11 | 0.52 (0.08, 3.01) | 0.47 | 0.62 (0.05, 5.72) | 0.69 | 0.74 (0.05, 8.43) | 0.82 |
| **26**<br>**A,B,C** | Rosetting | 1.71 (0.71, 4.14) | 0.23 | 3.88 (1.36, 10.99) | **0.01** | 3.64 (1.17, 11.26) | **0.02** |
|  | DC15 | 0.79 (0.14, 4.09) | 0.79 | 5.94 (0.9, 36.61) | 0.05 | 0.75 (0.04, 7.22) | 0.82 |
| **27**<br>**A,B,C** | Rosetting | 1.71 (0.71, 4.13) | 0.23 | 4.38 (1.55, 12.42) | **0.01** | 3.75 (1.2, 11.66) | **0.02** |
|  | DC16 | 0.83 (0.16, 3.72) | 0.81 | 0.32 (0.02, 2.69) | 0.35 | 0.49 (0.03, 4.43) | 0.57 |

### Group A-like *var* expression is independently associated with malaria mortality

Among the 966 included in this study 927 patients had a record of mortality status, of which, 58 died (**Table S1**). Among the 58 patients that died, 25 had overlapping syndromes, 25 had IC, 3 had SMA and 5 had moderate malaria while none died from either mild or RD (**Figure 7B**). We performed a logistic regression model with both group A-like *var* expression levels and Peripheral parasitaemia as explanatory variables with death as an outcome. Peripheral parasite density was not associated with child death [OR (95%CI): 1.00 (0.99, 1.00), p= 0.30]. However, death was predicted by group A-like *var* gene expression levels [OR (95%CI): 6.38 (2.28, 18.04), P= 0.0004], age and peripheral parasite density adjusted) (**Figure 7A**). The association was maintained even when the analysis was restricted to only children with impaired consciousness [OR (95%CI): 13.4(3.04, 63.), P = 0.0008, adjusted for age and peripheral parasite density). cys2 and cp2 were also associated with increased risk of mortality while gpB was associated with reduced risk of mortality. The rest of *var* subtypes showed no significant association with mortality (**Table S9**).

From the predicted *var* domains UPS groups and binding phenotypes, mortality was positively and significantly associated with DC16, DC15 and UpsA (from both cUPS and UpsML), EPCR-binding domains, while predicted DC9, UpsB, UpsC, and CD36 receptor binding domains showed significant negative associations with mortality (**Table S9**). We also tested whether VEH was associated with mortality. In a logistic regression model with death as the outcome, VEH showed a positive significant association with mortality [OR (95% CI): 7.67 (1.67, 30.49), P= 0.005 (**Table S9**).

**Figure 7:**
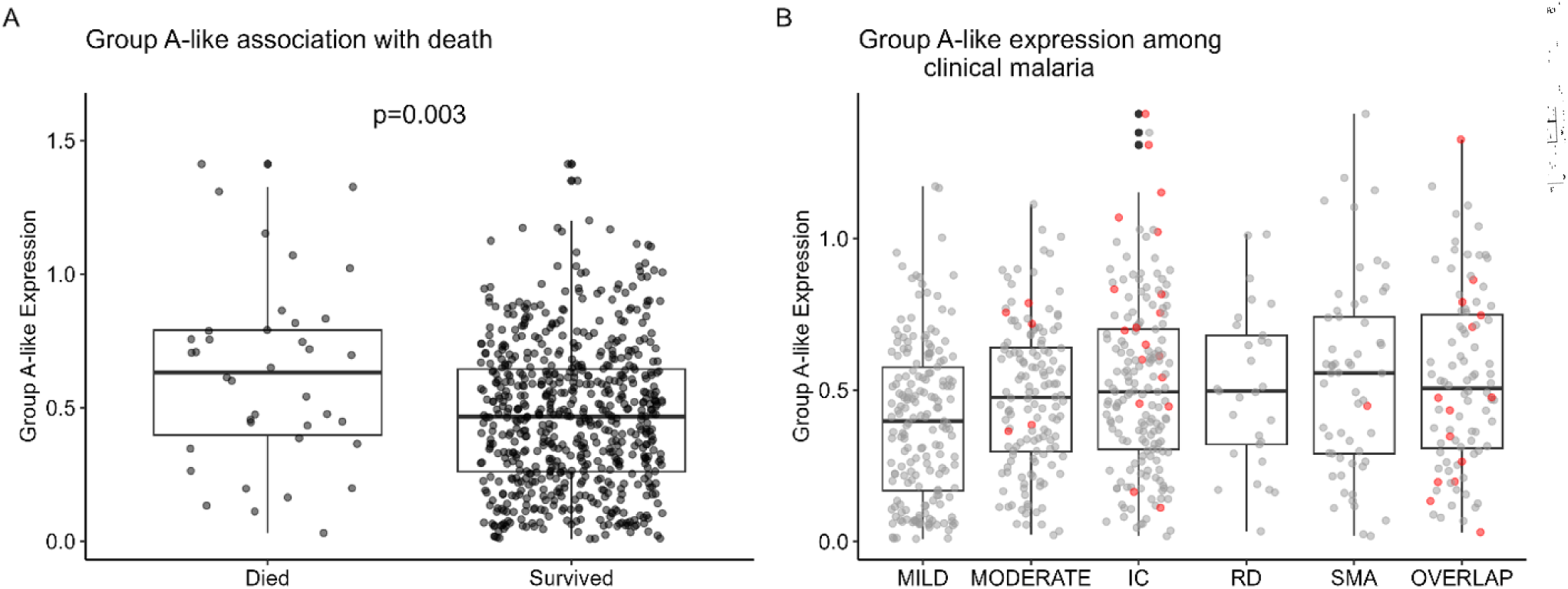
group A-like expression levels in relation to death. A) Group A–like expression in children who died (n = 58) versus those who survived (n = 871). P is a p value obtained from logistic regression model. B) Expression of Group A-like expression across clinical malaria syndromes; Red points indicate children who died. Number of deaths by syndrome: mild=0, moderate = 5, IC = 25, SMA = 3, overlap = 25, RD = 0. IC: impaired consciousness, RD: respiratory distress, SMA: severe malarial anaemia, overlap: mixed malaria infection with two or all the three severe malaria syndromes.

## Discussion

This study represents the largest analysis of *Plasmodium falciparum var* gene transcription across the major severe malaria syndromes associated with mortality: impaired consciousness (IC), respiratory distress (RD) and severe malarial anaemia (SMA) (Marsh et al., 1995; White, 2018; Kamau et al., 2022). Using DBLα-tag sequencing and RT-qPCR *var* datasets generated from parasite samples collected from 966 children recruited over an 18-year period in Kilifi, we show that severe malaria syndrome are associated with distinct patterns of parasite virulence. IC is consistently associated with increased expression of group A and DC8 *var* subtypes, RD is associated with rosetting and SMA is characterised by a relatively high homogeneous *var* expression profile, suggesting that the major severe malaria syndromes are associated with distinct parasite virulence strategies.

Our findings confirm and refine earlier observations from Kilifi and other African settings linking group A and DC8 *var* genes with pathogenesis of severe malaria, particularly cerebral malaria. Earlier studies identified associations between cp1/group A-like *var* genes and IC (Bull et al., 2005; Warimwe et al., 2009, 2012), DC8 and DC13 together with group A were enriched in severe disease (Lavstsen et al., 2012), and that EPCR- or dual EPCR-ICAM-1 binding *Pf*EMP1 variants mediate cytoadhesion of infected erythrocytes on human brain endothelial cells (Claessens et al., 2012; Avril et al., 2013; Turner et al., 2013). However, these studies were on a small sample size and largely compared severe to non-severe malaria or examined the pathogenic properties of specific *Pf*EMP1 variants *in vitro*. By directly comparing the major severe malaria syndromes within a single, well-characterised large cohort, we show that IC exhibits the most distinct and consistent *var* expression signature, with enrichment of group A and DC8 transcripts across both DBLα-tag sequencing, RT-qPCR and DBLα-tag sequencing prediction of UPS classification and receptor-binding phenotypes. Together with previous functional studies (Avril et al., 2013; Claessens et al., 2012; Turner et al., 2013), these findings support the concept that sequestration play a central pathogenic mechanism undeline IC.

The strong and consistent association of IC with EPCR-binding/group A *Pf*EMP1 variants also has important translational implications. Structural studies have demonstrated that CIDRα1-containing *Pf*EMP1 variants maintain conserved structural features despite extensive sequence diversity (Lau et al., 2015; Raghavan et al., 2023). Likewise, naturally acquired antibodies against recombinant DBLα domains belonging to the cp1 subgroup has been associated with protection against severe malaria (Tessema et al., 2019). Together, these findings support prioritising these virulence associated *Pf*EMP1 subsets as targets for vaccine development and therapeutic monoclonal antibodies aimed at preventing or disrupting sequestration of infected erythrocytes, thereby reducing the risk or severity of cerebral malaria (Avril et al., 2013; Reyes et al., 2024).

Unlike IC, RD was not associated with increased expression of any of the *var* subsets measured in this study, despite showing a clear association with the rosetting phenotype, consistent with previous observations (Warimwe et al., 2012). The persistence of the association between rosetting and RD after accounting for *var* expression suggests that variation in the *var* types measured in this study does not fully explain the link between rosetting and RD. Our RT-qPCR panel did not capture all PfEMP1 variants implicated in rosetting. Consistent with this possibility, predicted DC (11,15 and 16) which have recently been linked to rosetting (McLean et al., 2025), were correlated with rosetting in our study, but, were not significantly associated with RD. These findings suggest that additional parasite antigens may contribute to rosetting phenotype observed in RD. Therefore, we propose that future field studies should consider directly measuring DC11, DC15 and DC16 (McLean et al., 2025), as well as other variant surface antigens such as RIFINs (Goel et al., 2015), STEVORs (Niang et al., 2014) and SURFINs (Kanoi et al., 2020).

Rosetting is thought to promote parasite survival through at least two complementary mechanisms. First, by surrounding infected erythrocytes with uninfected erythrocytes, rosettes may physically shield the infected erythrocyte from immune recognition, thereby facilitating immune evasion (Moll et al., 2015). Second, the proximity of uninfected erythrocytes in a rosette may increase the efficiency of merozoite invasion immediately following schizont rupture, thereby enhancing parasite multiplication (Ribacke et al., 2013). The resulting increase in parasite biomass, which is consistent with our data (**Table S1A**), is likely to augment both parasite-derived lactate production and the host inflammatory response, with activated immune cells providing an additional source of lactate. Together, these processes could amplify hyperlactataemia and metabolic acidosis, the principal drivers of respiratory distress (English et al., 1996).

Although SMA showed some association with increased expression of group A *Pf*EMP1 and rosetting, its most distinguishing feature was the higher *var* expression homogeneity (a profile dominated by fewer variants). Previous studies have reported higher VEH in asymptomatic than clinical malaria, and proposed that this pattern reflect antibody-mediated selection pressure (Warimwe et al., 2013; Ceesay et al., 2025). In our cohort, however, SMA children were younger and had the lowest antibodies against infected erythrocyte among the malaria syndromes, suggesting that antibody-mediated selection is unlikely to explain higher VEH observed in SMA.

One possible explanation is that SMA may reflect more prolonged infections, during which within-host pressures select for a parasite population with a narrower *var* expression profile, dominated by group A or DC8 *var* genes. Although our cross-sectional design cannot establish infection duration, the longer reported fever duration among children with SMA prior to admission is consistent with this hypothesis. However, because fever duration was not significantly associated with VEH in our data, this interpretation remains speculative and will require further investigation in longitudinal studies. Nevertheless, prolonged infection could plausibly contribute both to the development of anaemia, through sustained red blood cell destruction including in the spleen and chronic inflammation that impair erythropoiesis (Phillips et al., 1986; Menendez et al., 2000; Lyke et al., 2004; White, 2018), and to the emergence of a more homogenous *var* transcriptional profile, through continued within-host positive selection.

The mortality patterns in our cohort underline the clinical relevance of group A-like *var* expression. Mortality was highest among children with IC, consistent with previous reports that impaired consciousness is a strong predictor of death among severe malaria syndromes (Marsh et al., 1995; Idro et al., 2007). Within this group, group A-like expression was associated both with IC and with death, corroborating previous observations that cp1/group A-like *var* subtypes are markers of life-threatening malaria (Warimwe et al., 2009, 2012). These findings suggest that group A *var* expression may have prognostic value, although its clinical utility will require validation in independent cohorts.

A key strength of this study is the combined use of DBLα-tag sequencing, RT-qPCR and newer bioinformatics classifiers, which together provide a more complete view of the *var* transcriptome than any single approach. DBLα-tag sequencing captures the overall breadth and relative composition of expressed *var* repertoires, whereas RT-qPCR allows precise quantification of clinically important *Pf*EMP1 subsets. The bioinformatics tools bridge these datasets by predicting UPS groups and domains from DBLα-tags, enabling direct comparison between sequence-based and primer-based measures and increasing confidence in the robustness of our findings. Considered together, these datasets revealed subtype syndrome associations that would likely have been missed using either method alone and showed that UPS group predictions and RT-qPCR classifications are largely complementary.

Nevertheless, several limitations warrant consideration. First, the analysis did not account for variations that may have been caused by parasite age at sampling and whole transcriptome data to indirectly stage the parasite was not available. However, the large sample size likely mitigated the confounding effects of variation in parasite stage. Second, our primer set targets a subset of known *Pf*EMP1 variants and does not capture all domain cassettes or binding phenotypes, and other multigene families implicated in rosetting and severe disease were not directly accessed. Third, host genetic and immunological factors may also influence *var* expression patterns and syndrome risk and have not been considered in this study.

In conclusion, the combined use of DBLα-tag sequencing, RT-qPCR and DBLα-tag-based prediction tools provide the most comprehensive characterisation to date of *P. falciparum var* gene expression across the major severe malaria syndromes. Our findings suggest that these syndromes are associated with distinct pathogenic mechanisms (graphical abstract). IC is characterised by preferential expression of EPCR or dual EPCR-ICAM-1 (DC8, and group A) *Pf*EMP1 binders, supporting sequestration as the dominant pathogenic mechanism, whereas RD appears more closely linked to downstream consequences of rosetting such as high parasitaemia and SMA to the consequences of chronicity as reflected by the relatively homogeneous *var* transcriptional profile. Together, these findings highlight the possibility of syndrome-specific parasite virulence mechanisms that may inform the development of targeted therapeutic interventions support the development of syndrome-specific interventions targeting distinct parasite virulence mechanisms. Future multi-omics studies integrating parasite transcription, adhesion phenotypes, host immunity and clinical phenotypes will be important for validating these mechanistic models and identifying new therapeutic targets.

## Data Availability

All packages used have been cited in the method. Data and R script used to perform the analysis is available at https://doi.org/10.7910/DVN/W4FFT9.

## Materials and Methods

### Study population

This study was conducted in Kilifi County, a malaria-endemic region located along the Kenyan coast. The patient samples analysed in this study have been previously described (Abdi et al., 2023). In brief, we analysed children who were slide-positive for *Plasmodium falciparum* and recruited between 1994 and 2012, a period spanning three distinct phases of malaria transmission in Kilifi; pre-decline, decline, and post-decline (Mogeni et al., 2016). Children were enrolled through Kilifi County Referral and Teaching hospital (KCRTH) admission surveillance (severe and moderate malaria) and from outpatient clinics (mild malaria). For the present analyses, we included only those with complete clinical data necessary for classification into the severe syndromes such as Blantyre coma score, deep-breathing and haemoglobin levels. Additionally, age and peripheral parasite density and the availability of *var* expression profiles measured by DBLα-tag sequencing or RT-qPCR were necessary for inclusion. This yielded 966 children; 898 with DBLα-tag data, 768 children with RT-qPCR data, and 702 with both, encompassing mild malaria, moderate, IC, RD, SMA and overlapping severe malaria syndromes.

### Definition of malaria syndromes

Severe malaria was defined as a hospital admission with any of the following; impaired consciousness (IC) defined using age-appropriate Blantyre coma score (BCS) thresholds (BCS of less than 5 for children over 8 months old or a BCS of less than 4 for children younger than 8 months) (Marsh et al., 1995; Warimwe et al., 2009), or respiratory distress (RD) (deep breathing) (English et al., 1996) or severe malarial anaemia (SMA) (hemoglobin < 5g/dl) (Marsh et al., 1995).

Non-severe Malaria (NSM) cases included malaria parasite-positive children managed at outpatient clinics, referred to as mild, and children admitted to hospital who were relatively stable and did not present with any of the three-life threatening severe syndromes described above, referred to as moderate(Marsh et al., 1995).

### Peripheral Blood Parasitaemia

Peripheral parasitaemia was quantified from Giemsa-stained thick and thin blood films following standard microscopy procedures. Parasite density was initially recorded as the number of infected erythrocytes per 500, 200, or 100 red blood cells (RBCs) examined. Parasitaemia per microlitre of blood was then calculated using World Health Organization (WHO) guidelines (WHO, 2010). Briefly, parasite density (parasites/µl) was derived using either:

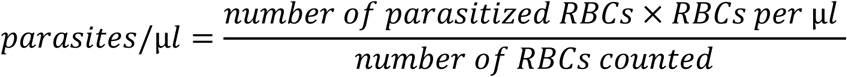

or, for counts performed against white blood cells (WBCs):

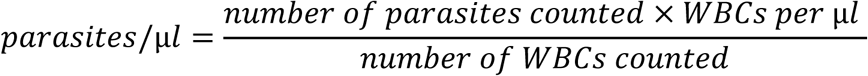

In instances where direct measurements of RBC or WBC concentrations were unavailable, standard assumptions of 5 × 10⁶ RBCs/µl and 8,000 WBCs/µl were applied, in accordance with WHO recommendations.

### *Pf*HRP2 Assay

Plasma concentrations of *P. falciparum* histidine-rich protein 2 (*Pf*HRP2) were measured using a sandwich ELISA as previously described (Abdi et al., 2023). In brief, Nunc MaxiSorp 96-well plates (Thermo Fisher Scientific) were coated overnight at 4°C with mouse anti-*Pf*HRP2 monoclonal antibody (MPFM-55A; MyBioscience) diluted in PBS to a final concentration of 0.9 µg/ml. Plates were washed three times with PBS containing 0.05% Tween-20 and subsequently blocked with PBS supplemented with 3% skimmed milk for 2 hr at room temperature (RT).

After an additional wash step, plasma samples and recombinant *Pf*HRP2 standards (MBS232321; MyBioscience), diluted in PBS with 2% BSA, were added in duplicate and incubated for 2 hr at RT. Plates were washed and incubated for 1 hr with HRP-conjugated mouse anti-*Pf*HRP2 detection antibody (MPFG-55P; MyBioscience) diluted in PBS/2% BSA to a final concentration of 0.2 µg/ml. After three washes, detection was achieved by incubating in o-phenylenediamine dihydrochloride (OPD) substrate for 15 min, and reactions were stopped with 2M H₂SO₄. Optical density (OD) was read at 490 nm using a BioTek Synergy4 plate reader. The ODs where log-transform for further analysis.

### RNA and cDNA preparation

The method for RNA extraction and cDNA preparation from the samples used in this study was described previously (Abdi et al., 2016; Warimwe et al., 2012). Briefly, the RBC fraction was enriched by depleting the buffy coat from whole blood samples. The enriched RBC fraction was resuspended in TRIzol (Invitrogen, Paisley, United Kingdom) and stored at −80°C. After thawing, the total RNA was extracted by chloroform phase separation and GlycoBlue/isoamyl alcohol precipitation. Pellets were washed with 75% ethanol, briefly air-dried, and re-suspended in RNAsecure. Residual genomic DNA was removed by DNase I treatment (DNase-Free, Ambion) and inactivated with the Ambion DNase Inactivation Reagent according to the manufacturer’s instructions. cDNA was prepared by reverse transcribing RNA using reverse transcriptase (Invitrogen Superscript III). For each cDNA reaction, a negative control reaction was performed in the absence of reverse transcriptase to ensure that all contaminating DNA had been removed by DNAse pre-treatment. No sample showed any amplification in these controls.

### *Var* sequencing and classification

The expressed sequence tag (EST) sequencing approach used to generate this dataset has been previously described (Abdi et al., 2016; Warimwe et al., 2009). In summary, degenerate primers (Bull et al., 2005), were used to amplify a region within the DBLα-tag domain that is highly conserved across most *var* genes. Two sequencing methods were employed: capillary sequencing and the 454-pyrosequencing platform.

For the capillary sequencing approach, amplified DBLα-tags were cloned into the TOPO®-TA cloning vector and transformed into E. coli. From each sample, up to 96 colonies were picked and sequenced individually. For the 454 platform, the amplicons were barcoded, multiplexed, and sequenced using high-throughput 454 sequencing (Kivisi et al., 2019). The median number of reads obtained per sample was 2833 (range: 20-8964). The Sample with more than 20 reads were included in the analysis.

Sequences obtained from each sample were classified and counted as previously described (Bull et al., 2005; Warimwe et al., 2009). Proportional expression was calculated as the proportion of sequences within each sample assigned to a specific sequence group. DBLα-tag sequences were characterised using complementary classification schemes based on number of cysteine (cys) residues, positions of limited variability (PoLV), and sharing of polymorphic sequence blocks (bs).

Based on the number of cysteine residues, DBLα-tag sequences were classified into three groups: cys2 (two cysteine residues), cys4 (four cysteine residues), and cysX (one, three, five, or six cysteine residues). Combining cysteine number with the presence or absence of the conserved PoLV motifs MFK and REY generated six cysteine/PoLV (cp) groups (cp1-cp6; **Table 1**). All known cp1 sequences belong to group A *var* genes, although not all group A *var* genes are classified as cp1 (Bull et al., 2007).

The second classification scheme was based on block sharing (bs), which groups DBLα-tag sequences according to networks of recombining polymorphic sequence blocks. bs1 and bs2 are the major block-sharing groups, with bs1 enriched for group A *var* genes. Sequences belonging to bs1 and containing two cysteine residues (bs1_cys2) are referred to as group A-like sequences (Warimwe et al., 2009).

The cp and bs classifications are complementary rather than equivalent. In this study, cys2, cp1, cp2, cp3, and bs1_cys2 (group A-like) were included in the analysis because of their previously reported association with severe malaria (Warimwe et al., 2009).

### Upstream sequence (UPS) group and domain prediction

DBLα-tag contigs generated from patient samples were used to predict the upstream sequence (UPS) group and the PfEMP1 domain. The upstream sequence group was predicted using cUPS and upsML with default parameters. cUPS (https://github.com/qianfeng2/cUps) is a probabilistic classifier that assigns DBLα-tag sequences to the most likely var upstream promoter group (UPSA, UPSB, or UPSC) and reports classification probabilities for each assignment (Tan et al., 2025). upsML (https://github.com/sii-scRNA-Seq/upsML) is a machine learning-based classifier for predicting a var upstream group from DBLα-tag sequences (Pangilinan et al., 2026). Domain composition was predicted using Varia with default parameters (https://github.com/GCJMackenzie/Varia). Varia predicts PfEMP1 domain composition by comparing query sequence tags to database genes based on sequence similarity (Mackenzie et al., 2022). Predicted domain compositions were then used to assign binding phenotype (Rask et al., 2010; Jespersen et al., 2016; McLean et al., 2025), and domain-cassette classifications based on defined domain motifs arrangement (Rask et al., 2010) (Table S2). Predicted UPS group, domain type, domain-cassette assignments, and binding phenotypes were summarised at the sample level, and proportions were calculated for downstream analyses.

### RT-PCR *var* transcription profiles

The qRT-PCR primers used to determine *var* transcript profile data in this study were as used by Abdi et al, (Abdi et al., 2016) and described previously (Rottmann et al., 2006; Rask et al., 2010; Lavstsen et al., 2012). Data were generated using primers that showed differential expression between severe and non-severe malaria in previous studies (Lavstsen et al., 2012). These included four primer sets targeting DC8 (named dc8-1, dc8-2, dc8-3, dc8-4), one primer set for each of DC13 (dc13), and DC9 (dc9), two primer sets targeting the majority of group A *var* genes (gpA1 and gpA2). Group B and C *var* gene transcription was quantified using primer sets b1 and c2 respectively. The names and sequences of the original primers are shown in **Table S4.** In addition, we included a primer targeting *Pf*sir2a, which has been associated with dysregulation of *var* gene expression (Merrick et al., 2012, Abdi et al., 2016).

Levels of *var* gene transcription were normalized relative to the housekeeping gene *seryl tRNA synthetase* and *fructose bisphosphate aldolase*. The arbitrary transcript unit (Tu_s_) was calculated using the formula (Tu_s_ = 2^(5−Δct)^) where Δct is the averaged change in cycle threshold of *var* gene relative to housekeeping genes (Lavstsen et al., 2012; Abdi et al., 2016).

### *Var* gene expression homogeneity (VEH) Calculation

To quantify the degree of dominance among *var* gene transcripts expressed by each parasite isolate in a sample, we assessed *Var* gene expression homogeneity (VEH). VEH reflects the extent to which a small number of *var* gene sequences dominate an isolate’s expression profile (Warimwe et al., 2013).

We employed Simpson’s Index of Diversity to capture the overall homogeneity of the *var* expression repertoire (Simpson, 1949; Warimwe et al., 2013). For each sample, we calculated Simpson’s Index (λ) as the sum of the squares of proportions of all distinct *var* gene expressed in each sample:

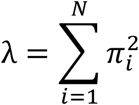

Where; *π_i_* is the proportion of *var* gene i among N total *var* genes counted in a sample. The index ranges from 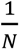 (indicating maximal diversity and minimal homogeneity) to 1 (indicating complete dominance by a single sequence and thus maximal VEH).

### Rosetting Assay

Parasite isolates directly from patients’ blood were matured *ex vivo* to mature trophozoite stage before cryopreservation for 24 hrs. On the day of assay, cryopreserved trophozoites -infected erythrocytes were thawed (Kinyanjui et al., 2004). Following thawing, Trophozoite-infected erythrocytes were then adjusted to 4% haematocrit in RPMI 1640 medium (Gibco) supplemented with 5μg/ml acridine orange (Sigma-Aldrich). For each isolate, 9.5μL of this suspension was mixed with 2.5μL of non-immune AB serum in a U-bottomed 96-well plate and rotated vertically at room temperature for 30 minutes. The sample was transferred onto a microscope slide, covered with an 18mm × 18mm coverslip, and examined under a fluorescence microscope (Nikon Eclipse 80i, Japan) at 40× magnification. Rosette frequency was determined as the percentage of 200 trophozoite-infected erythrocytes binding to two or more uninfected erythrocytes. Scoring was conducted without knowledge of the clinical category of the patient from whom each isolate originated.

### Antibody Recognition of trophozoite-infected erythrocytes (α-IE Assay)

Total IgG binding to the surface of trophozoite-infected erythrocytes (IEs) was quantified by flow cytometry, as described previously (Kivisi et al., 2019). Cryopreserved mature trophozoite-stage IEs from two heterologous parasite isolates, A4U and 11019, were thawed on the day of the assay (Kinyanjui et al., 2004), and used directly. A4U is a long-term laboratory-adapted isolate derived from IT and selected to predominantly express the group B PfEMP1 A4VAR/ITVAR14/PfIT_130005200, which binds ICAM-1 and CD36 (Mackintosh et al., 2008). Isolate 11019 is a recently culture-adapted clinical isolate from a child with cerebral malaria that displays a rosetting phenotype and predominantly expresses the group A DC11 PfEMP1 PFKE10VAR_R1/g54 (McLean et al., 2025).

For each assay well, 11μL of an infected erythrocyte suspension containing mature trophozoite-stage IEs (prepared from A4U and 11019 at approximately 1-2% parasitaemia in 1×PBS/0.5%BSA and ethidium bromide) was incubated with 1µL of each child’s acute plasma (final plasma dilution 1:12) for 30 min at room temperature. This was followed by two washes in 200µL 1×PBS/0.5%BSA and incubated for 30 min in the dark with 50µL fluorescein isothiocyanate (FITC)-conjugated sheep anti-human IgG (cat# AF004) diluted 1:50 in 1×PBS/0.5%BSA. The samples were finally washed as before and then analysed on BD FACSCanto II cytometer.

Antibody reactivity was expressed as the median fluorescence intensity (MFI) of infected erythrocytes after subtraction of the corresponding uninfected erythrocyte signal. To account for non-specific antibody binding, the mean MFI obtained with plasma from four non-exposed European donors included on each assay plate was subtracted from the corrected MFI of each child’s sample. Negative background-adjusted MFI values were set to zero and retained in downstream analyses, indicating no detectable antibody recognition above background. Plasma from a semi-immune Kilifi adult was included as a positive control. The gating strategy used is presented Figure S5.

### Statistical analysis

We selected samples with both RT-qPCR and DBLα-tag sequence transcript data for analysis. RT-qPCR-derived *var* transcript expression values were log-transformed, while proportions of DBLα-tag-defined *var* subtypes were arcsine square root transformed (Warimwe et al., 2009). To identify associations between *var* gene expression and severe malaria syndromes (IC, RD, and SMA), binary variables of the severe syndromes were created, and non-severe cases were considered as controls. We fitted multiple linear regression models using stats R core package (R Core Team, 2026). The model followed the general structure below in equation 1:

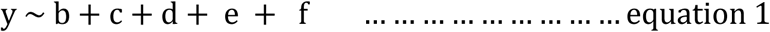

Where:

*y* represents specific *var* gene defined by RT-qPCR primer sets or DBLα-tag subtype, *b* represents binary representation where all children with IC syndrome were assigned 1 and those without it were assigned 0, *c* represents binary representation where children with RD syndrome were assigned 1 and those without were assigned 0, *d*; represents binary representation where all children with SMA syndrome were assigned 1 and those without were assigned 0. *e*; represents age of the participant in months while *f* represent log transformed values of peripheral blood parasite density. This analysis was repeated without children with overlapping syndromes to identify syndrome-specific *var* gene associations.

Model outputs included beta coefficient of estimate with 95% confidence interval (β (95%CI) and corresponding p values. To correct for multiple testing, p values were adjusted using Benjamin-Hochberg false discovery rate (FDR). Associations were considered statistically significant when adjusted p value was less than 0.05. Positive beta (β) values indicate that the variable was positively associated with the malaria syndrome relative to the control while the negative β values indicated inverse association. Results are presented as forest plots and summary tables.

We compared VEH scores across pure clinical presentations using two-sided Wilcoxon rank-sum tests to determine the relationship between syndrome and *var* expression homogeneity. To explore determinants of VEH, we assessed associations with host factors (α-IE, fever duration before admission and febrile temperature) and parasite factors (*Pf*sir2a, peripheral blood parasite density and RT-qPCR *var* gene transcript expression levels) using Spearman’s rank correlation. We report correlation coefficients (rho) and p values, and visualize group differences and associations using ggplot2 in R.

Rosette frequency was quantified as the proportion of infected erythrocytes forming rosettes in ex vivo parasite isolates. Differences between severe malaria (n = 145) and non-severe malaria (n = 128) were assessed using the Wilcoxon rank-sum test. Syndrome-specific comparisons were then performed using pairwise Wilcoxon rank-sum tests for children with and without impaired consciousness (IC), respiratory distress (RD), and severe malaria anaemia (SMA). To assess the independent association between rosetting and each severe malaria syndrome while accounting for overlapping clinical presentations, linear regression models were fitted with arcsine square root-transformed rosette frequency as the outcome and IC, RD, and SMA as predictors, adjusting for age and peripheral parasite density.

Correlation between rosette frequency and *var* gene expression was assessed using Spearman’s rank correlation coefficient. To estimate 95% confidence intervals (95% CIs) for the correlation coefficients, we applied non-parametric bootstrapping with 5000 replicates using boot R package (Canty & Ripley, 2016). This was followed by evaluating the relationship between rosetting and individual severe malaria syndromes. We constructed a linear regression model as in equation 1 and replaced *y* with rosette frequency. To test whether the relationship between rosetting and the severe syndromes is independent of *var* expression, we conducted logistic regression models considering each of the syndromes as outcome with rosette frequency as a predictor while adjusting for expression levels of *var* gene subtypes.

To identify parasite factors associated with mortality, we fitted binary logistic regression models with death (yes/no) as the outcome. We first ran univariable models for each *var* transcript measure and for parasite density separately and then fitted multivariable models including the *var* measure and parasite density together to estimate the effect of each on death while adjusting for age in all models. Odds ratios (ORs) with 95% CIs are reported.

All analyses were conducted in R version 4.6.1 (R Core Team, 2026). Forest plots of effect sizes from linear and logistic regression models were generated using ggplot2 (Wickham, 2016). Data manipulation was performed using dplyr and tidyverse packages (Wickham et al., 2026), and model tidying was implemented with broom.

## Ethical approval

This study obtained ethical approval from the Scientific Ethics Review Unit of the Kenya Medical Research Institute (KEMRI/SERU/3149) and written informed consent was obtained from parents or guardians of the study participants.

## Author contribution

**Henry Ndugwa** Conceptualization, Investigation, methodology, Formal analysis, Visualization, Writing – original draft, review and editing,

**Michelle Muthui** methodology, Writing–review and editing,

**J. Alexandra Rowe** Writing –review and editing

**Kinyanjui SM**, Project administration, Supervision, Conceptualization, analysis, Writing - original draft, review and editing

**Cheryl Andisi Kivisi** Project administration Supervision, Conceptualization. Methodology, analysis, Writing – original draft, review and editing

**Abdirahman I. Abdi** Conceptualization, Supervision, formal analysis, Writing – original draft, review and editing, All authors contributed to the interpretation of the analyses and revised the draft manuscript.

## Acknowledgement and funding

This work was supported by Wellcome Trust Awards: 209289/Z/17/Z (to Abdirahman I. Abdi). Henry Ndugwa is fully funded by the Science for Africa Foundation to the Developing Excellence in Leadership, Training and Science in Africa (DELTAS Africa) programme [DEL-22-012] with support from Wellcome Trust and the UK Foreign, Commonwealth & Development Office and is part of the EDCPT2 programme supported by the European Union.

## Supplementary materials

**Figure S1:**
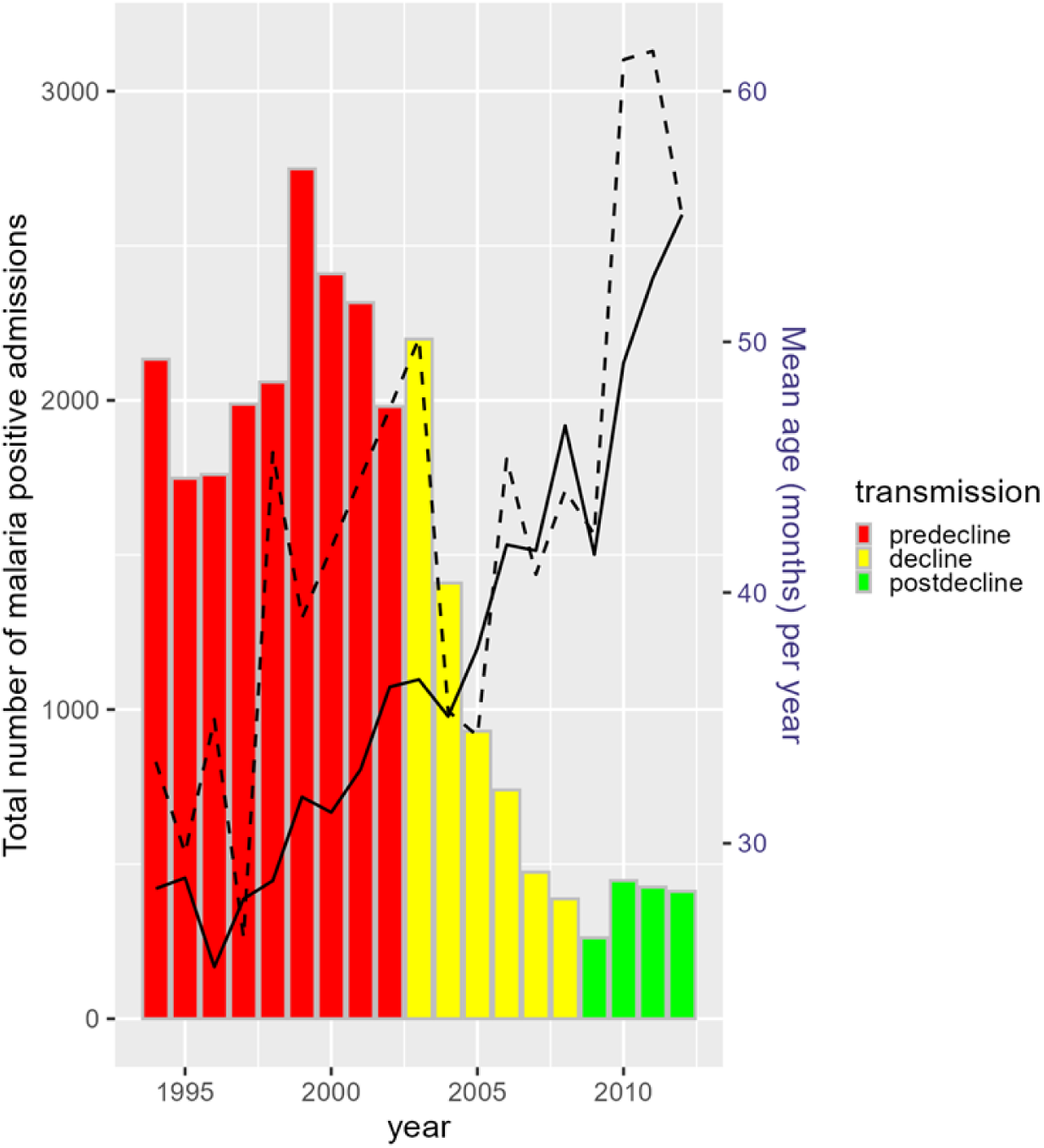
Total malaria admissions and mean patient age per year. Number of patients per year (left axis), mean patient age per year (right axis). The solid black line is the average patient age of total admission, the dashed line is the average patient age in this study

**Figure S2.**
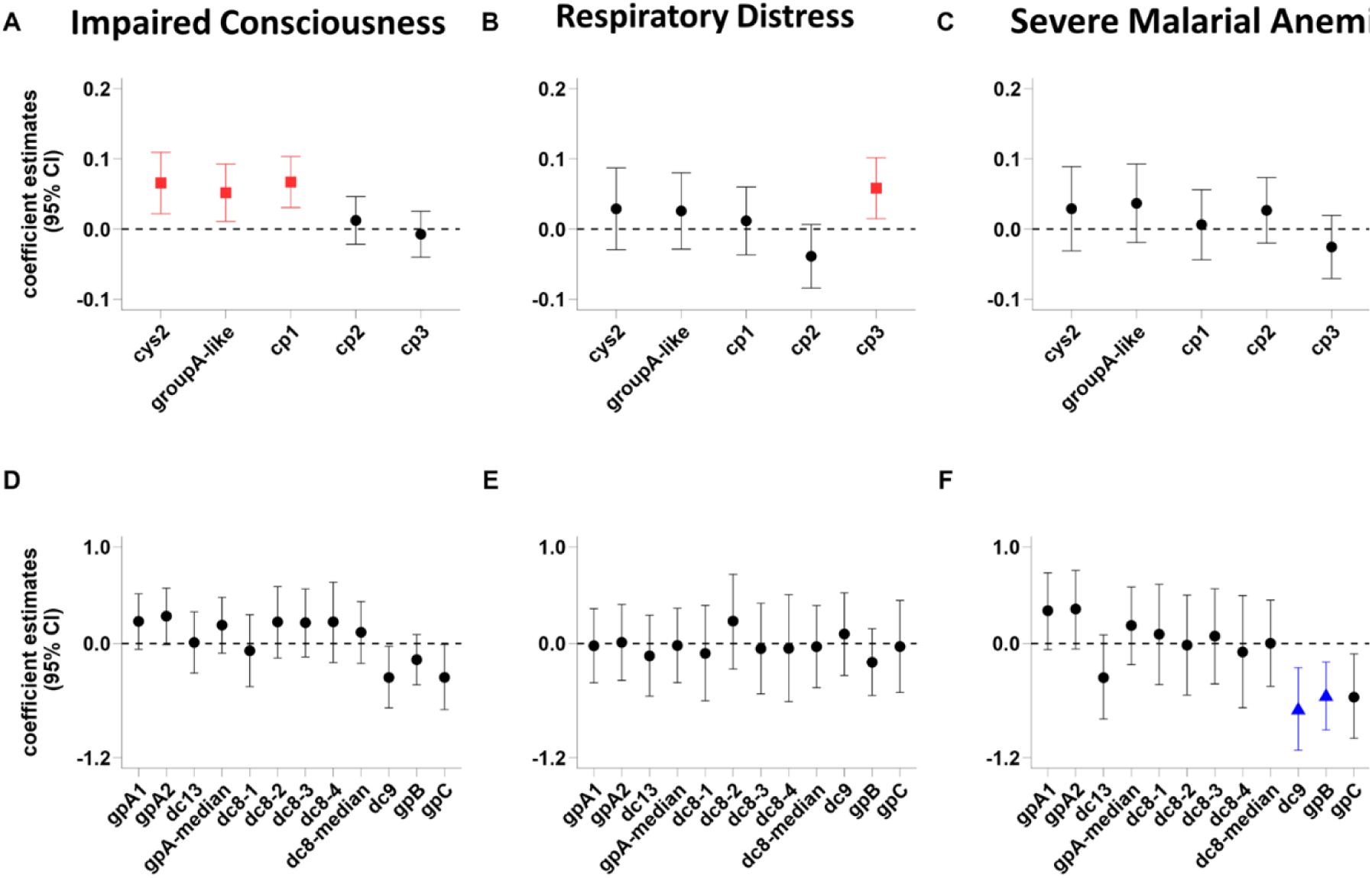
Associations between *var* gene expression and severe malaria syndromes without previously analysed data. **A-C).** Coefficients (95% CI) from linear regression models examining associations between the expression of cys2 and its subgroups (group A-like, cp1, cp2, and cp3) and the severe malaria syndromes impaired consciousness (IC), respiratory distress (RD), and severe malarial anaemia (SMA), adjusting for age and parasitaemia. **D-F).** Coefficients (95% CI) from analogous models examining associations between transcript levels of gpA1-gpC and the same severe malaria syndromes, also adjusted for age and parasitaemia. Points represent regression coefficients and error bars indicate 95% confidence intervals. Sample sizes were: non-severe malaria (NSM), n = 304; IC, n = 207; RD, n = 32; SMA, n = 67; and overlapping syndromes, n = 106.

**Figure S3.**
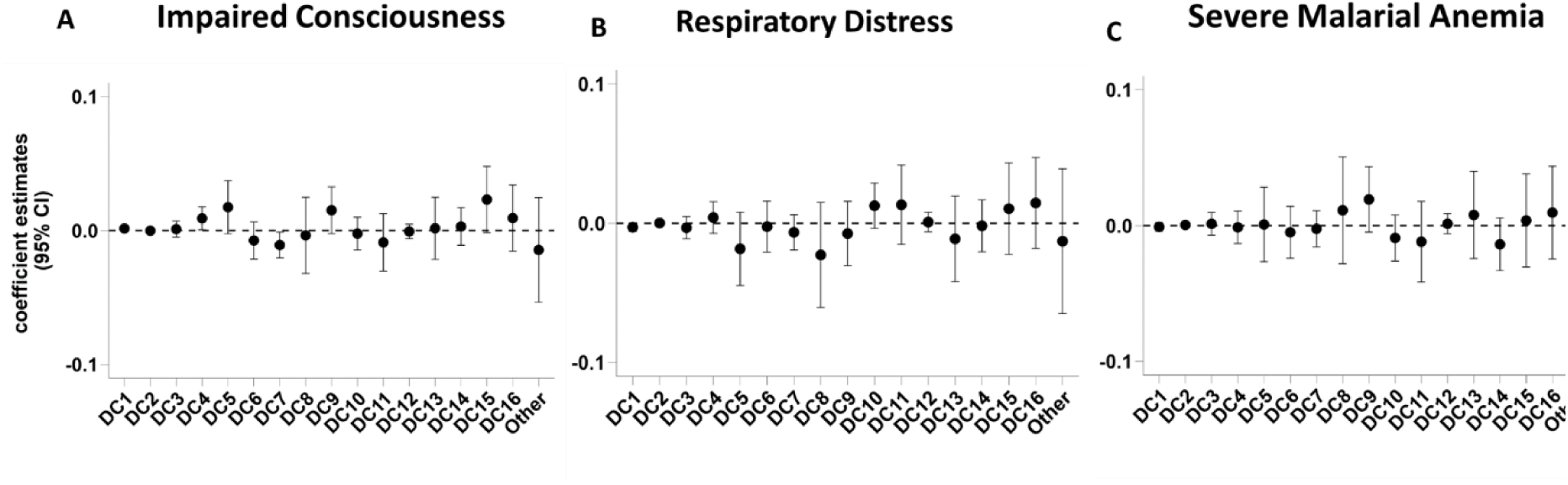
Associations between predicted var gene domain cassette and severe malaria syndromes. Coefficient estimates (95% CI) from multiple linear regression models examining associations between domain cassette (DC)-level *var* gene expression and severe malaria syndromes: impaired consciousness (IC, left), respiratory distress (RD, middle) and severe malarial anaemia (SMA, right); For each model, the outcome variable was the proportion of DBLα-tag sequences assigned to a given DC category, while the explanatory variable was the corresponding clinical syndrome. All models were adjusted for age and parasite density. Points represent regression coefficients and error bars indicate 95% confidence intervals. The horizontal dashed line represents the null effect (coefficient = 0).

**Figure S4.**
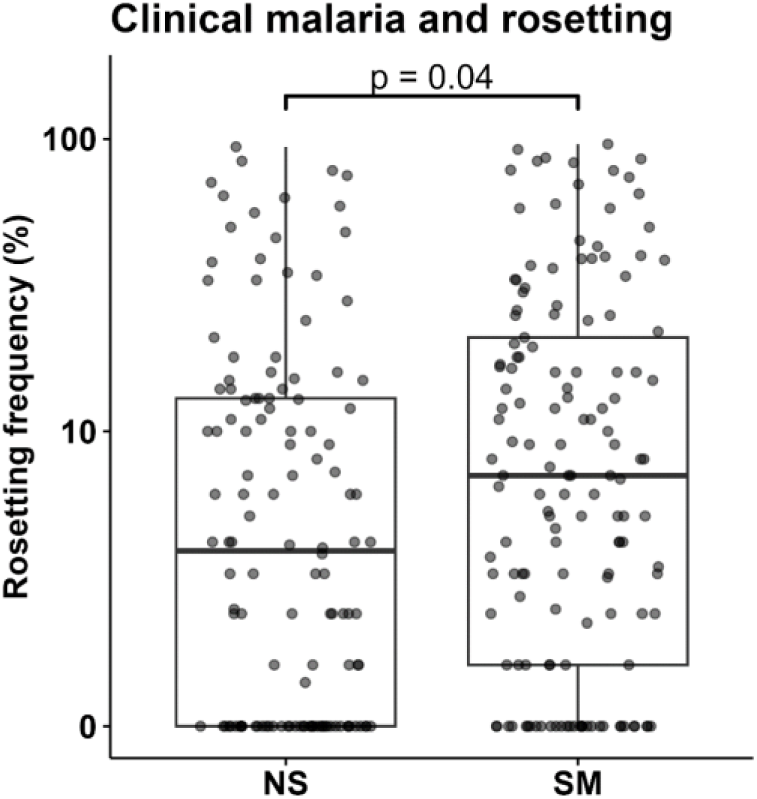
Association between clinical malaria severity and rosetting frequency. Rosette frequencies were compared between children with non-severe malaria (NS) and severe malaria (SM). Boxplots show the median, interquartile range, and overall distribution of rosette frequencies, with individual observations overlaid as points. Children with severe malaria exhibited significantly higher rosette frequencies than those with non-severe malaria (Wilcoxon rank-sum test, *P* = 0.04).

**Figure S5.**
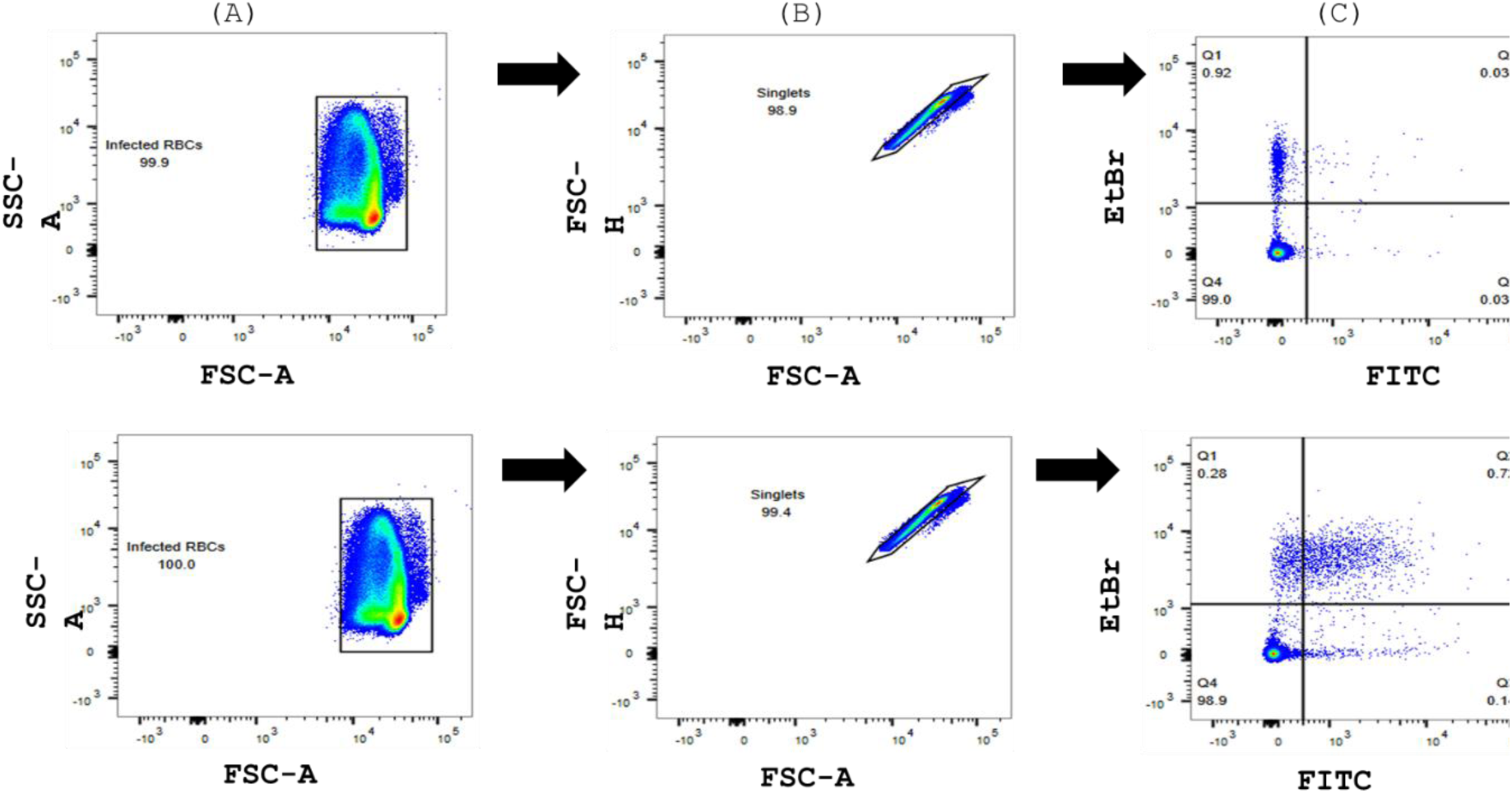
Gating strategy used to quantify plasma IgG binding to mature trophozoite-stage infected erythrocytes. Upper and the lower panels represent examples of the reactivity of plasma samples from unexposed and malaria exposed donors respectively on mature pigmented trophozoite-stage infected erythrocytes of A4U and 11019 isolates. **(A).** Cells were first gated based on forward scatter (FSC-A) and side scatter (SSC-A) characteristics to exclude debris, then gated for **(B)** singlets (FSC-H vs FSC-A). **(C).** Infected erythrocytes were identified by positive ethidium bromide (EtBr) staining. Surface-reactive antibodies were detected using FITC-conjugated secondary antibodies, and variant surface antigen (VSA) reactivity was quantified as the difference between the median FITC fluorescence intensity of infected erythrocytes and that of uninfected erythrocytes. Representative plots illustrate the low reactivity observed for plasma from an unexposed donor and the higher reactivity observed for plasma from a malaria-exposed donor.

**Table S1A:** Patient characteristics (DBL-tag Data). Significant p values are in bold

| Syndrome | Age (months)<br>(Median [IQR]) | Pvalue | Parasitemia<br>[Median[IQR]]<br>(x1000) | Pvalue | Mortality<br>(%) | Pvalue |
| --- | --- | --- | --- | --- | --- | --- |
| **Non-Severe Malaria** (n=398) | 43[24–68] | Ref | 180.00[39.36–363.60] | Ref | 5 (1%) | Ref |
| **Severe Malaria** (n= 500) | 37[22–55] | **<0.001** | 180.00[14.76–461.50] | 0.305 | 46 (10%) | **<0.001** |
| **IC** (n=249) | 40[26–61] | 0.153 | 149.80[11.36–417.40] | 0.073 | 21 (9%) | **<0.001** |
| **RD** (n=41) | 48[31–60] | 0.647 | 320.00[48.00–721.60] | **0.052** | 0 (0%) | 1 |
| **SMA** (n=76) | 30[13–41] | **<0.001** | 167.08[20.02–328.20] | 0.241 | 3 (4%) | 0.122 |
| **IC/SMA** (n=28) | 28[18–41] | **0.002** | 98.80[15.20–363.00] | 0.19 | 2 (7%) | 0.071 |
| **RD/IC** (n=79) | 36[25–54] | **0.048** | 253.20[24.45–531.80] | 0.383 | 16 (20%) | **<0.001** |
| **SMA/RD** (n=10) | 34[30–40] | 0.271 | 199.20[71.10–351.20] | 0.997 | 0 (0%) | 1 |
| **IC/SMA/RD** (n=17) | 21[12–42] | **0.003** | 100.80[29.15–352.80] | 0.678 | 4 (27%) | **<0.001** |

**Table S1B:** Patient characteristics (RT-qPCR Data). Significant p values are in bold

| Syndrome | Age (months)<br>(Median [IQR]) | Pvalue | Parasitemia<br>[Median[IQR]] (x1000) | Pvalue | Mortality<br>(%) | Pvalue |
| --- | --- | --- | --- | --- | --- | --- |
| **Non-Severe Malaria** (n=359) | 46[26–73] | Ref | 170.00[38.67–350.00] | Ref | 5 (1%) | Ref |
| **Severe Malaria**(n=409) | 36[22–50] | **<0.001** | 190.40[25.52–432.00] | 0.494 | 42 (11%) | **<0.001** |
| **IC** (n=206) | 39[26–58] | **0.002** | 150.60[21.29–414.40] | 0.639 | 24 (12%) | **<0.001** |
| **RD** (n=29) | 26[12–38] | 0.722 | 272.00[23.54–500.00] | 0.169 | 0 (0%) | 1 |
| **SMA** (n=59) | 26[12–38] | **<0.001** | 190.00[67.00–407.40] | 0.377 | 1 (2%) | 1 |
| **IC/SMA** (n=20) | 27[18–36] | **<0.001** | 126.60[35.78–363.00] | 0.93 | 2 (10%) | **0.047** |
| **RD/IC** (n=70) | 36[24–51] | **0.003** | 257.00[44.80–509.40] | 0.086 | 12 (17%) | **<0.001** |
| **SMA/RD** (n=6) | 32[28–39] | 0.18 | 252.40[171.90–351.20] | 0.395 | 0 (0%) | 1 |
| **IC/SMA/RD** (n=19) | 24[15–36] | **<0.001** | 97.20[13.19–361.80] | 0.405 | 3 (18%) | **0.005** |

**Table S2:**
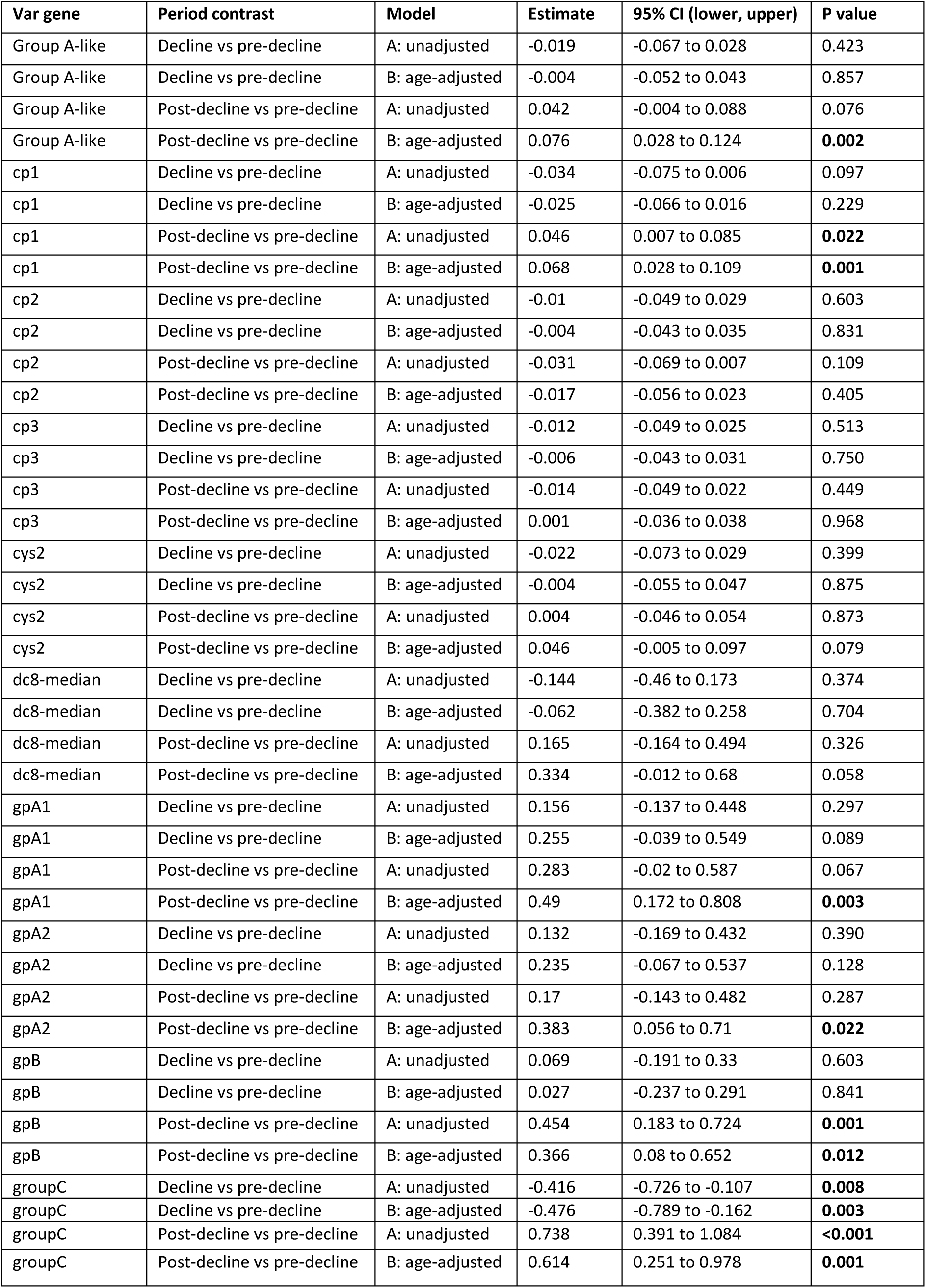
Associations between transmission period, age, and var gene expression. Linear regression models were fitted for each var subtype with transmission as the main predictor, Model A includes transmission period only (unadjusted), and Model B additionally adjusts for age in months

**Table S3:** Multiple linear regression estimates for associations between *var* variants (cys2, group A-like, cp1, cp2, and cp3) and severe malaria syndromes.

| Primary analysis with all the syndromes |  |  |  |  |  |  |  |  |  |
| --- | --- | --- | --- | --- | --- | --- | --- | --- | --- |
| Var subtype | Impaired Consciousness |  |  | Respiratory Distress |  |  | Severe Malarial Anaemia |  |  |
|  | Estimate (95% CI) | P.Value | Adjusted P.value | Estimate (95% CI) | P.Value | Adjusted P.value | Estimate (95% CI) | P.Value | Adjusted P.value |
| **cys2** | 0.09 (0.05, 0.13) | **<0.0001** | **<0.0001** | 0.03 (-0.02, 0.08) | 0.2606 | 0.2722 | 0.06 (-0.00, 0.11) | 0.0533 | 0.1331 |
| **groupA-like** | 0.08 (0.04, 0.11) | **<0.0001** | **<0.0001** | 0.03 (-0.02, 0.08) | 0.2639 | 0.2722 | 0.06 (0.01, 0.12) | **0.0151** | 0.0753 |
| **cp1** | 0.08 (0.05, 0.11) | **<0.0001** | **<0.0001** | 0.02 (-0.02, 0.07) | 0.2722 | 0.2722 | 0.02 (-0.02, 0.07) | 0.3039 | 0.3799 |
| **cp2** | 0.02 (-0.01, 0.05) | 0.1703 | 0.2129 | -0.03 (-0.07, 0.01) | 0.149 | 0.2722 | 0.03 (-0.02, 0.07) | 0.219 | 0.365 |
| **cp3** | 0.00 (-0.03, 0.03) | 0.9234 | 0.9234 | 0.04 (0.00, 0.08) | 0.0455 | 0.2273 | -0.01 (-0.05, 0.03) | 0.6513 | 0.6513 |
| Analysis without overlapping severe malaria syndromes |  |  |  |  |  |  |  |  |  |
| **cys2** | 0.09 (0.04, 0.13) | **0.0003** | **0.0004** | 0.05 (-0.04, 0.15) | 0.2664 | 0.444 | 0.05 (-0.02, 0.13) | 0.1648 | 0.2746 |
| **groupA-like** | 0.09 (0.04, 0.13) | **0.0001** | **0.0002** | 0.07 (-0.02, 0.15) | 0.1288 | 0.3357 | 0.08 (0.01, 0.14) | **0.0289** | 0.1444 |
| **cp1** | 0.08 (0.04, 0.12) | **<0.0001** | **0.0002** | 0.02 (-0.06, 0.09) | 0.6089 | 0.7611 | 0.01 (-0.05, 0.07) | 0.6783 | 0.6783 |
| **cp2** | 0.03 (-0.01, 0.07) | 0.1047 | 0.1309 | 0.00 (-0.07, 0.08) | 0.952 | 0.952 | 0.04 (-0.01, 0.10) | 0.1453 | 0.2746 |
| **cp3** | -0.00 (-0.04, 0.03) | 0.8301 | 0.8301 | 0.05 (-0.02, 0.12) | 0.1343 | 0.3357 | -0.02 (-0.07, 0.04) | 0.5422 | 0.6777 |

**Table S4:**
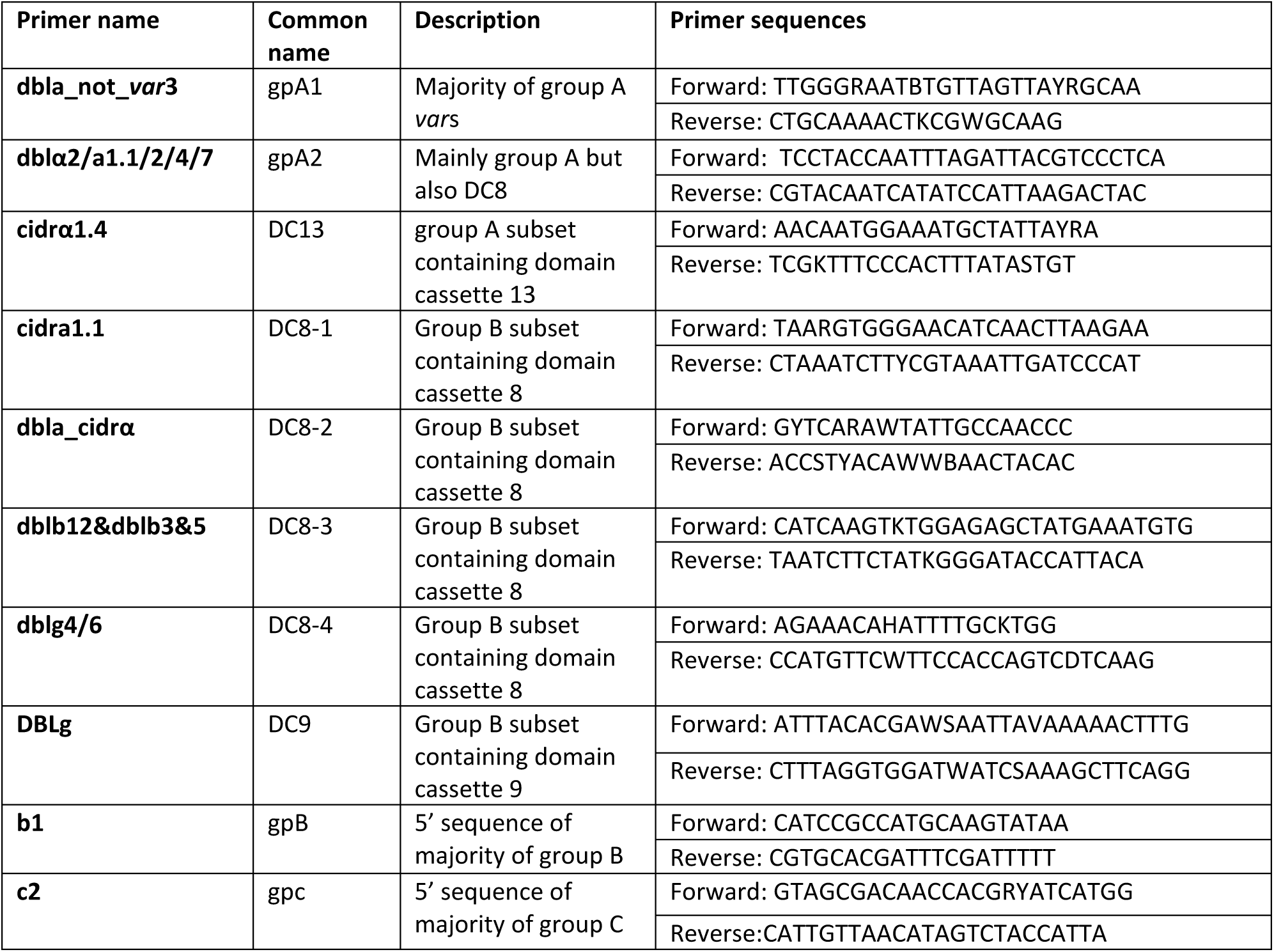
Description of Primers used in quantifying var genes.

**Table S5:** Multiple linear regression estimates for associations between *var* variants (gpA1-gpC) and severe malaria syndromes; Impaired Consciousness.

| Primary analysis with all the syndromes |  |  |  |  |  |  |  |  |  |
| --- | --- | --- | --- | --- | --- | --- | --- | --- | --- |
| Var subtype | Impaired Consciousness |  |  | Respiratory Distress |  |  | Severe Malarial Anaemia |  |  |
|  | Estimate (95% CI) | P.Value | Adjusted P.value | Estimate (95% CI) | P.Value | Adjusted P.value | Estimate (95% CI) | P.Value | Adjusted P.value |
| gpA1 | 0.38 (0.12, 0.63) | **0.0035** | **0.042** | 0.02 (-0.32, 0.35) | 0.9276 | 0.9276 | 0.28 (-0.08, 0.64) | 0.1221 | 0.2921 |
| gpA2 | 0.33 (0.07, 0.59) | **0.012** | **0.0481** | 0.05 (-0.29, 0.39) | 0.7669 | 0.9165 | 0.39 (0.03, 0.76) | **0.0329** | 0.1316 |
| dc13 | 0.17 (-0.09, 0.44) | 0.1969 | 0.2626 | -0.07 (-0.42, 0.28) | 0.6868 | 0.9165 | -0.30 (-0.67, 0.07) | 0.1107 | 0.2921 |
| gpA-median | 0.30 (0.05, 0.56) | **0.0195** | 0.0585 | 0.03 (-0.30, 0.37) | 0.8401 | 0.9165 | 0.24 (-0.12, 0.59) | 0.1947 | 0.2921 |
| dc8-1 | 0.22 (-0.11, 0.54) | 0.1923 | 0.2626 | -0.26 (-0.69, 0.17) | 0.2331 | 0.9165 | 0.34 (-0.12, 0.79) | 0.15 | 0.2921 |
| dc8-2 | 0.35 (0.03, 0.67) | **0.0313** | 0.0626 | 0.08 (-0.34, 0.50) | 0.7172 | 0.9165 | 0.07 (-0.38, 0.51) | 0.7737 | 0.7737 |
| dc8-3 | 0.40 (0.10, 0.70) | **0.0088** | **0.0481** | -0.10 (-0.49, 0.30) | 0.6361 | 0.9165 | 0.16 (-0.26, 0.58) | 0.4591 | 0.6121 |
| dc8-4 | 0.28 (-0.07, 0.62) | 0.1125 | 0.1929 | 0.05 (-0.41, 0.51) | 0.83 | 0.9165 | -0.10 (-0.59, 0.38) | 0.6822 | 0.7442 |
| dc8-median | 0.31 (0.03, 0.58) | **0.0281** | 0.0626 | -0.11 (-0.48, 0.25) | 0.5449 | 0.9165 | 0.11 (-0.28, 0.49) | 0.5816 | 0.6979 |
| dc9 | 0.03 (-0.25, 0.30) | 0.8492 | 0.88 | 0.11 (-0.26, 0.47) | 0.5624 | 0.9165 | -0.51 (-0.89, -0.12) | **0.0099** | 0.0592 |
| gpB | -0.02 (-0.24, 0.21) | 0.88 | 0.88 | -0.19 (-0.50, 0.11) | 0.2051 | 0.9165 | -0.50 (-0.82, -0.18) | **0.0022** | **0.0259** |
| gpC | 0.06 (-0.22, 0.35) | 0.6692 | 0.8031 | -0.07 (-0.46, 0.32) | 0.7236 | 0.9165 | -0.26 (-0.64, 0.13) | 0.1877 | 0.2921 |
| Analysis without overlapping severe malaria syndromes |  |  |  |  |  |  |  |  |  |
| gpA1 | 0.52 (0.22, 0.81) | **0.001** | **0.002** | 0.71 (0.07, 1.35) | 0.0296 | 0.0711 | 0.52 (0.04, 0.99) | **0.0335** | 0.0671 |
| gpA2 | 0.55 (0.25, 0.84) | **0.0003** | **0.0017** | 1.01 (0.37, 1.66) | **0.002** | **0.024** | 0.77 (0.29, 1.24) | **0.0017** | **0.0206** |
| dc13 | 0.19 (-0.11, 0.49) | 0.214 | 0.2854 | 0.03 (-0.62, 0.68) | 0.929 | 0.929 | -0.07 (-0.55, 0.41) | 0.7785 | 0.7785 |
| gpA-median | 0.50 (0.21, 0.79) | **0.001** | **0.002** | 0.87 (0.23, 1.51) | **0.0079** | **0.0472** | 0.63 (0.15, 1.11) | **0.0098** | **0.042** |
| dc8-1 | 0.34 (-0.04, 0.71) | 0.0822 | 0.1157 | 0.28 (-0.54, 1.09) | 0.5041 | 0.6049 | 0.75 (0.15, 1.35) | **0.0148** | **0.0445** |
| dc8-2 | 0.62 (0.26, 0.97) | **0.001** | **0.002** | 0.91 (0.13, 1.69) | **0.0216** | 0.0647 | 0.60 (0.02, 1.18) | **0.0417** | 0.0715 |
| dc8-3 | 0.68 (0.34, 1.02) | **0.0001** | **0.001** | 0.90 (0.17, 1.64) | 0.0159 | 0.0634 | 0.71 (0.17, 1.26) | **0.0105** | **0.042** |
| dc8-4 | 0.43 (0.03, 0.83) | **0.0333** | 0.0571 | 0.50 (-0.36, 1.37) | 0.2561 | 0.4391 | 0.23 (-0.41, 0.87) | 0.4834 | 0.5801 |
| dc8-median | 0.53 (0.21, 0.84) | **0.001** | **0.002** | 0.64 (-0.04, 1.33) | 0.0636 | 0.1273 | 0.56 (0.05, 1.06) | **0.0319** | 0.0671 |
| dc9 | 0.10 (-0.21, 0.41) | 0.5285 | 0.5766 | 0.33 (-0.36, 1.02) | 0.3445 | 0.5167 | -0.40 (-0.91, 0.11) | 0.1229 | 0.1638 |
| gpB | 0.04 (-0.22, 0.30) | 0.781 | 0.781 | -0.09 (-0.65, 0.48) | 0.7631 | 0.8324 | -0.36 (-0.78, 0.06) | 0.0923 | 0.1385 |
| gpC | 0.17 (-0.17, 0.51) | 0.3173 | 0.3807 | 0.23 (-0.43, 0.90) | 0.4919 | 0.6049 | -0.13 (-0.64, 0.38) | 0.6168 | 0.6729 |

**Table S6A:** Description of Domain Cassette classification.

| Domain cassette | Domain motif detected | Associated phenotype | Reference |
| --- | --- | --- | --- |
| DC1 | DBLα1.1/1.4–CIDRα1.2/1.3–DBLβ1.1/11–DBLγ1/15–DBLε1 ... DBLγ8–DBLζ1/2–DBLε5 | unknown | (Rask et al., 2010) |
| DC2 | DBLpam1–DBLpam2–CIDRpam | placental sequestration (CSA) | (Salanti et al., 2003; Rask et al., 2010; Clausen et al., 2012) |
| DC3 | DBLα1.3–DBLε8 | unknown | (Rask et al., 2010) |
| DC4 | DBLα1.1/1.4–CIDRα1.6–DBLβ3 | ICAM-1 binding DBLβ3; group A | (Bengtsson et al., 2013) |
| DC5 | DBLγ12–DBLδ5–CIDRβ3/4–DBLβ7/9 | PECAM-1 binding | (Rask et al., 2010; Turner et al., 2013) |
| DC6 | DBLγ14–DBLζ5–DBLε4 | unknown | (Rask et al., 2010) |
| DC7 | DBLε2–DBLε7–DBLε3 | unknown | (Rask et al., 2010) |
| DC8* | DBLα2–CIDRα1.1/1.8–DBLβ12–DBLγ4/6 | EPCR binding (CIDRα1.1) | (Rask et al., 2010; Turner et al., 2013; Lau et al., 2015; McLean et al., 2025) |
| DC9 | DBLγ3–DBLζ4 | unknown | (Rask et al., 2010) |
| DC10 | DBLζ6–DBLε9 | unknown | (Rask et al., 2010) |
| DC11 | DBLα1.8–CIDRβ/$\gamma$/$\delta$ | Rosetting and IgM-Fc binding | (Rask et al., 2010; McLean et al., 2025) |
| DC12 | DBLζ3–DBLε12 | unknown | (Rask et al., 2010) |
| DC13 | DBLα1.7–CIDRα1.4 | EPCR binding/ ICAM-1-binding DBLβ | (Turner et al., 2013; Lau et al., 2015) |
| DC14 | DBLα0.6–CIDRα3.1–DBLβ5 | unknown | (Rask et al., 2010) |
| DC15 | DBLα1.2–CIDRα1.5 | Rosetting | (Rask et al., 2010; McLean et al., 2025) |
| DC16 | DBLα1.5/1.6–CIDRδ/$\gamma$ | Rosetting | (Rask et al., 2010; McLean et al., 2025) |

**\*some, but not all, DC8 variants are EPCR binders.**

**Table S6B:** Description of Predicted domain binding phenotype classification.

| Priority | Predicted binding phenotype | Domain arrangement/criterion | Reference |
| --- | --- | --- | --- |
| 1 | CSA | DBL<sub>pam1</sub>–DBL<sub>pam2</sub>–CIDR<sub>pam</sub> | (Salanti et al., 2003; Clausen et al., 2012) |
| 2 | EPCR-ICAM- 1 dual binding | DBLα1.4/1.7–CIDR α1.4/1.6-DBLβ3 | (Avril et al., 2016; Lennartz et al., 2017) |
| 3 | ICAM-1 | DBLα0.3/0.4–CIDR α2-6-DBLβ5 | (Bengtsson et al., 2013; Lennartz et al., 2017) |
| 4 | PECAM-1 | DBLγ12–DBLδ5–CIDRβ3/4–DBLβ7/9 | (Berger et al., 2013) |
| 5 | EPCR | CIDRα1.1/1.4–1.8 | (Turner et al., 2013; Lau et al., 2015) |
| 6 | CD36 | CIDRα2–6 | (Kraemer & Smith, 2003; Robinson et al., 2003) |
| 7 | gC1qR | DBLβ12 | (Magallón-Tejada et al., 2016) |

**NB:** Predicted binding phenotypes were assigned according to the order shown above. Where a sequence matched multiple criteria, it was assigned to the highest-priority phenotype.

**Table S7:** Expression of Pfsir2a among clinical syndromes. P values are derived from Wilcoxon rank-sum test

| **group** | **p value** |
| --- | --- |
| IC vs SMA | **0.026** |
| Mild vs IC | 0.063 |
| RD vs SMA | 0.088 |
| Moderate vs IC | 0.166 |
| Mild vs RD | 0.196 |
| Moderate vs RD | 0.257 |
| Moderate vs SMA | 0.257 |
| Mild vs SMA | 0.406 |
| IC vs RD | 0.658 |
| Mild vs Moderate | 0.702 |

**Table S8:**
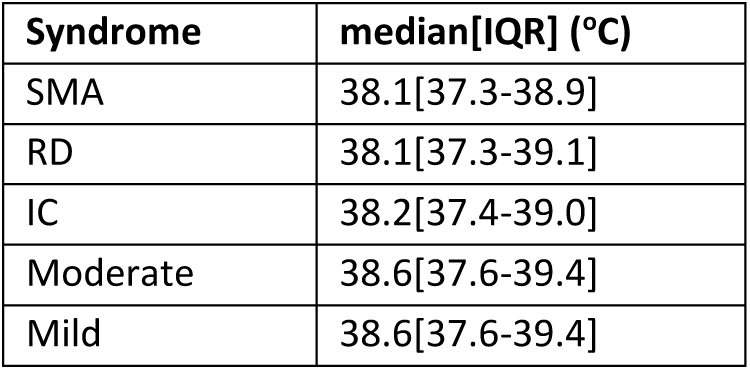
The median body temperature within each syndrome at admission.

| **Syndrome** | **median[IQR] (°C)** |
| --- | --- |
| SMA | 38.1[37.3-38.9] |
| RD | 38.1[37.3-39.1] |
| IC | 38.2[37.4-39.0] |
| Moderate | 38.6[37.6-39.4] |
| Mild | 38.6[37.6-39.4] |

**Table S9:** Association of var gene expression, and mortality. p value<0.05 are highlighted (bold)

| **Var subtypes** | **OR (95%CI)** | **p value** | **Data type** |
| --- | --- | --- | --- |
| cys2 | 4.58 ( 1.72 , 12.4 ) | **0.002** | DBLα-tag |
| groupA-like | 6.38 ( 2.28 , 18.04 ) | **0.0004** | DBLα-tag |
| cp1 | 1.81 ( 0.56 , 5.59 ) | 0.309 | DBLα-tag |
| cp2 | 3.08 ( 1.00 , 8.83 ) | **0.042** | DBLα-tag |
| cp3 | 1.92 ( 0.50 , 6.76 ) | 0.325 | DBLα-tag |
| gpA1 | 1.18 ( 0.98 , 1.44 ) | 0.086 | RT-qPCR |
| gpA2 | 1.16 ( 0.97 , 1.41 ) | 0.112 | RT-qPCR |
| dc13 | 0.95 ( 0.79 , 1.12 ) | 0.563 | RT-qPCR |
| gpA-median | 1.13 ( 0.94 , 1.36 ) | 0.214 | RT-qPCR |
| dc8-1 | 1.12 ( 0.98 , 1.3 ) | 0.107 | RT-qPCR |
| dc8-2 | 1.09 ( 0.95 , 1.26 ) | 0.226 | RT-qPCR |
| dc8-3 | 1.15 ( 0.98 , 1.36 ) | 0.085 | RT-qPCR |
| dc8-4 | 1.06 ( 0.93 , 1.19 ) | 0.363 | RT-qPCR |
| dc8-median | 1.11 ( 0.94 , 1.33 ) | 0.218 | RT-qPCR |
| dc9 | 0.9 ( 0.76 , 1.07 ) | 0.239 | RT-qPCR |
| gpB | 0.77 ( 0.64 , 0.94 ) | **0.008** | RT-qPCR |
| gpC | 0.82 ( 0.67 , 1.00 ) | 0.059 | RT-qPCR |
| VEH | 7.67 ( 1.67 , 30.49 ) | **0.005** | VEH |
| UPSA | 4.6 ( 1.69 , 12.37 ) | **0.003** | cUPS |
| UPSB | 0.3 ( 0.08 , 1.18 ) | 0.083 | cUPS |
| UPSC | 0.05 ( 0.01 , 0.28 ) | **0.001** | cUPS |
| EPCR | 3.09 ( 1.23 , 7.76 ) | **0.016** | Predicted binding |
| ICAM1 | 0.06 ( 0.00 , 1.78 ) | 0.161 | Predicted binding |
| ICAM1+EPCR | 0.17 ( 0 , 3.46 ) | 0.336 | Predicted binding |
| PECAM1 | 1.53 ( 0.11 , 10.16 ) | 0.7 | Predicted binding |
| CD36 | 0.23 ( 0.08 , 0.63 ) | **0.004** | Predicted binding |
| Unknown | 2.56 ( 0.13 , 24.17 ) | 0.468 | Predicted binding |
| DC4 | 1.32 ( 0 , 43.78 ) | 0.903 | Predicted DC |
| DC5 | 2.54 ( 0.35 , 12.58 ) | 0.229 | Predicted DC |
| DC6 | 0.21 ( 0 , 4.72 ) | 0.402 | Predicted DC |
| DC7 | 0.49 ( 0 , 24.35 ) | 0.773 | Predicted DC |
| DC8 | 0.23 ( 0.04 , 1.1 ) | 0.087 | Predicted DC |
| DC9 | 0.02 ( 0 , 0.46 ) | **0.034** | Predicted DC |
| DC10 | 0.36 ( 0 , 9.34 ) | 0.605 | Predicted DC |
| DC11 | 0.8 ( 0.1 , 4.61 ) | 0.822 | Predicted DC |
| DC12 | 0.22 ( 0 , 175.16 ) | 0.715 | Predicted DC |
| DC13 | 1.98 ( 0.36 , 8.53 ) | 0.39 | Predicted DC |
| DC15 | 4.61 ( 1.2 , 15.34 ) | **0.017** | Predicted DC |
| DC16 | 5.23 ( 1.35 , 17.97 ) | **0.011** | Predicted DC |
| Other-dcs | 0.39 ( 0.14 , 1.12 ) | 0.081 | Predicted DC |

